# A Turn in the Road: Real-life Driving Differences in Obstructive Sleep Apnoea and Excessive Daytime Sleepiness

**DOI:** 10.1101/2025.10.14.25337986

**Authors:** Kieran O. Lee, Tristan A. Bekinschtein, Ian E. Smith

## Abstract

**Background:** A driver with untreated obstructive sleep apnoea (OSA) is at an increased relative risk of crashing their vehicle. Nonetheless, the chance of any individual being involved in a collision remains low and it is not clear how we can identify which drivers with OSA are most at risk. Since motion detection is used by insurers to assess risky driving, we sought to characterise how driving changes with OSA and daytime sleepiness in real life.

**Methods:** Ninety-one recruited participants (mean age 51 [22 to 77] years; 35% female; mean ESS 12.5, mean ODI 15/h) completed 7,834 journeys (mean duration 15.6 minutes), yielding 508,544 driving events. Participants were recruited at referral to our sleep service and installed a smartphone app that recorded driving data, at least until diagnostic outcomes were known. Data were analysed for driving events of braking, accelerating and turning using linear mixed-effects models against participant data including age, sex, BMI, Epworth Sleepiness Scale (ESS), oxygen desaturation index (ODI), time into trip and journey duration.

**Findings:** ESS and ODI combined showed all clinical groups robustly differed from non-OSA / non-EDS, [ODI<5 / ESS<11] in turning behaviour. Younger age was linked to different, potentially riskier behaviour, while ESS and ODI, when analysed independently, showed no robust association with identifiable differences in driving.

**Interpretation:** Smartphone telematics can distinguish people with self-reported sleepiness and OSA from non-OSA / non-EDS individuals, via turning behaviour. This supports shifting emphasis from diagnosis and self-report towards objective behavioural indicators of impairment. Combining telematics with cognitive, physiological and/or subjective measures could yield practical criteria to improve crash-risk assessments and provide objective support for decisions around driving cessation or continuance.

**Funding:** This research was supported by funds from the Royal Papworth Charity and Consciousness and Cognition Lab.

**Research in context:** *Evidence before this study:* We searched Google Scholar and PubMed from inception to September 1^st^ 2025, with no language restrictions, using terms related to ‘obstructive sleep apnoea’, ‘sleepiness’, ‘driving’, ‘crash’, and ‘telematics’. Existing literature consistently links OSA with increased crash risk. Almost all reviewed studies used cohort, database reviews, self-report or driving simulator designs, which lack accurate real-world risk prediction at subgroup and individual level. This prominent issue has led to high profile organisations including the European Respiratory Society, The National Sleep Foundation and The Lancet to call for better research into detection of at-risk sleepy drivers.

*Added value of this study:* In a prospective clinical cohort at referral, we captured smartphone passive telematics (Insights, Sentiance) across 7,834 journeys, generating 508,544 events from 91 adults. Using linear mixed-effects models, we show that turning behaviour differentiated participants with combined sleepiness and OSA from those with neither (ODI <5 / ESS <11), whereas ESS or ODI alone were not independently associated with identifiable differences in braking/acceleration profiles. This study demonstrates the feasibility and signal of smartphone-derived features to help phenotyping driving risk within routine OSA pathways.

*Implications of all the available evidence:* Smartphone telematics could improve current risk assessments for drivers with suspected or confirmed OSA, offering scalable, low-cost, passively collected markers that align with real-world behaviour. If externally validated, manoeuvre-specific metrics (e.g., turning) may help triage patients for further evaluation, tailor driving advice or restrictions, and provide objective support for decisions regarding driving cessation or continuance. Next steps include independent replication, integrating these data with cognitive, physiological and subjective dynamics, linkage to hard outcomes (near-misses and crashes), assessment of fairness and privacy, and development of clinically usable thresholds suitable for regulatory and occupational settings.

## Introduction

Untreated OSA can cause sleep fragmentation and nocturnal hypoxia, and is associated with excessive daytime sleepiness (EDS), impaired cognition, and increased risk of a motor vehicle collision. Large meta-analyses reviewing OSA and driving behaviour have estimated that collision risk is between 1.21 and 4.89 times higher than in the general population (1,2), causing significant loss of life, injury and financial cost. These findings are supported by government and insurance records (3–5), emergency department attendance (6) and self-report measures (7,8). However, the chance of any individual being involved in a collision is low (2) and some drivers display resilience to impairment (9). This makes revoking rights to drive problematic.

Previous research has assessed clinical markers of OSA, and demographic characteristics to establish if high-risk individuals can be identified with readily available information, but results have been inconsistent. While EDS is intuitively the cause of excess driving risk, and there is some evidence that degree of sleepiness is related to impairment, in a meta-analysis it was reported that in half of the studies reviewed there was no significant relationship between subjective daytime sleepiness and crash risk (1). In another study, performance on multiple sleep latency tests and maintenance of wakefulness tests were associated with driving simulator task performance (10) but these tests are not readily available.

OSA severity, in terms of both the apnoea-hypopnoea index (AHI), and oxygen saturation (SpO_2_), have also been shown to be related to risk in some studies (8,11–13). However, others did not find a relationship (14–18). Again looking to the published meta-analysis, while the majority of 19 reviewed studies with a control group showed an increased risk of crashes in people diagnosed with sleep apnoea less than half showed an association between risk and the severity of OSA (1).

Driving simulators that have been used to predict driving risk in people with OSA, have the advantage of being accessible to people when they have been instructed not to drive and provide reproducible test conditions. One such study combined OSA severity and EDS, and found that together they explained less than 25% of the variance in driving performance (19). However, the results are not obviously applicable to real-world driving. For example, women have shown worse performance on driving simulator tasks (18,20,21) but this is not reflected in the real-world crash risk and could be related to testing procedures favouring men. The European Respiratory Society issued a statement on sleep apnoea, sleeping and driving risk in 2021, where they stress the strong need for good quality studies to detect sleepy drivers and that performance on driving simulators does not predict accidents or performance in real-life, and that future research should investigate outcomes in real-life situations (22). Other organisations such as the National Sleep Foundation (23) and recent editorials (24,25), have called for similar collective efforts.

Developments in passive telematics data acquisition via smartphones have made it feasible to collect real-world driving performance data in discrete populations. Such data have been used commercially to more accurately profile drivers, adjust insurance premiums, and improve business modelling (26). One published study assessed 40 million journeys undertaken by Uber drivers, and found that tipping was greater for journeys with data reflective of a smoother driving profile (27). In a clinical population, 131 apparently cognitively normal adults underwent screening for Alzheimer’s disease and recorded driving telematics. Driving performance was related to severity of preclinical biomarkers for cognitive impairment (28). In another study data from 120,000 journeys were collected by 96 community-dwelling older adults some with sleep apnoea. Showed that an eight-point increase in apnoea-hypopnoea index (AHI) was associated with a 1.27 times increase in the chance of an adverse driving event (29).

In the current study, we test if real-world driving data, collected via telematics methods, are related to clinical markers of OSA, and demographics, at the peri-diagnosis stage of OSA across an otherwise unselected population referred to a sleep clinic for investigation of possible sleep apnoea.

## Methods

### Study Design and Participants

This study used an observational design to measure the real-world driving behaviour of patients attending Royal Papworth Hospital NHS Foundation Trust, Sleep Centre, Cambridge, UK, for OSA screening. Between August 2023 and August 2024, patients were invited to take part via a letter with a QR code linked to the study information sheet, and a screening questionnaire which could be completed by interested potential participants. Exclusion criteria included no driver’s licence, not currently driving, being under the age of eighteen years and use of CPAP (continuous positive airway pressure) within the last 6 months. Following confirmation of eligibility, willing participants provided informed consent remotely via the online platform DocuSign.

Enrolled participants downloaded the mobile phone application ‘Insights’ (developed by Sentiance) supported by a member of the study team. They were asked to drive as usual, recording data from set-up of the app until at least when the results of their diagnostic tests were available. Some patients diagnosed with OSA were then advised not to drive pending the initiation of treatment as per our clinical practice. Patients started on treatment were asked to continue or resume data collection. See Figure 1. Diagnostic and demographic data were extracted from the hospital’s electronic patient record system. OSA severity was based on the Oxygen Desaturation Index (ODI) from home oximetry, or Apnoea-Hypopnoea Index (AHI) if the patient underwent respiratory polygraphy or polysomnography. In our sample 73 (80.2%) of the participants had only overnight oximetry, and for simplicity we grouped ODI and AHI, under the label ODI. Self-reported sleepiness was assessed with the Epworth Sleepiness Scale (ESS). Ethical approval for the study was granted by the UK Health Research Authority (IRAS Project ID: 314762).

**Figure 1:**
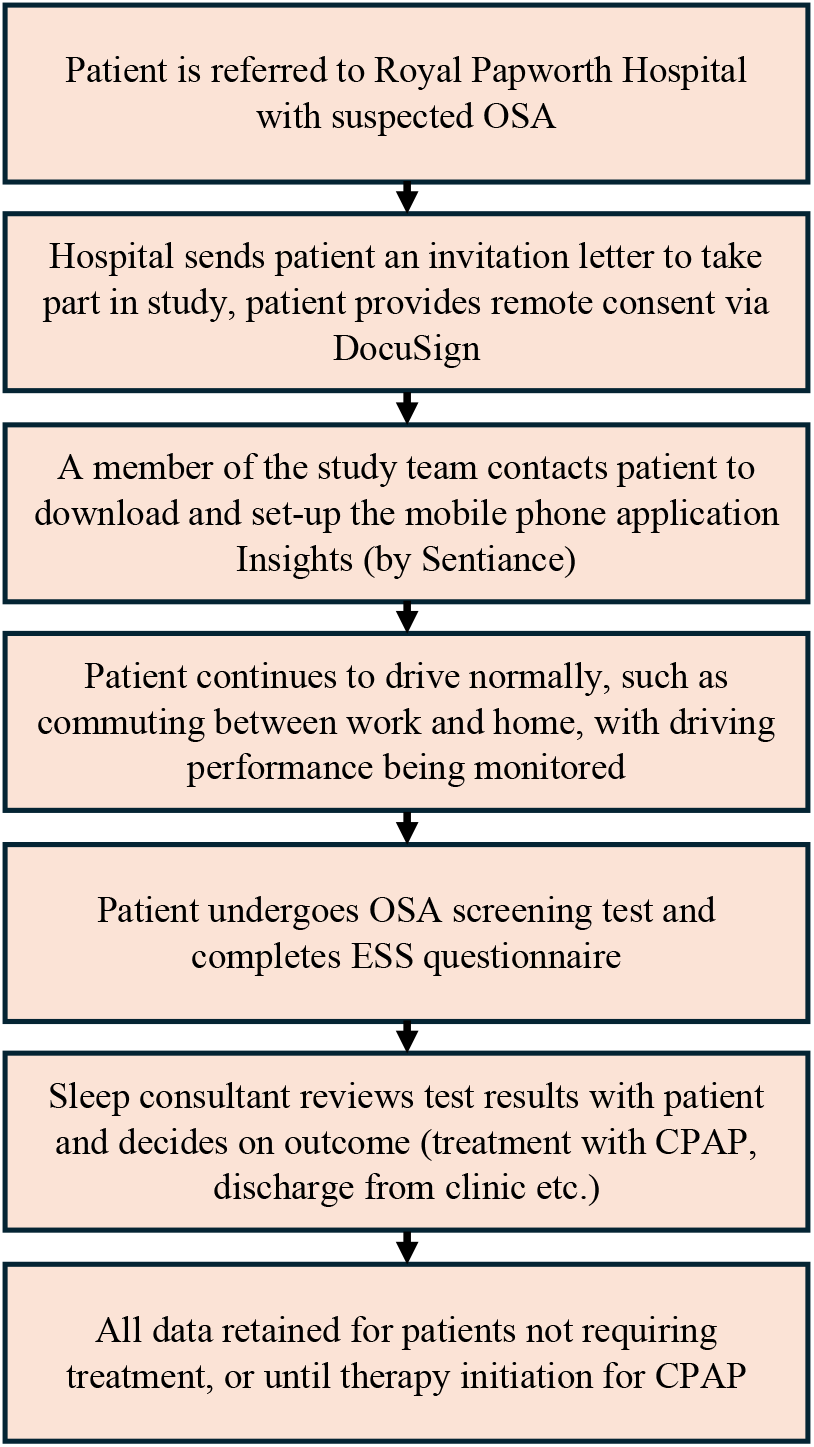
Flow-chart of study procedure.

### Driving Performance Monitoring Equipment

The Insights application collects data from the gyroscope and accelerometer, which are included in most modern mobile phones. The data are analysed to produce specific timestamped events of braking, accelerating and turning which contain data for magnitude and duration of these events. If events exceeded predetermined limits they were categorised as ‘significant’. The frequency of significant events per minute of a journey is reported. We utilised nine measures of driving performance. Six individual event measures, and three frequency measures: brake magnitude (BM), brake duration (BD), significant brake frequency (SBF), acceleration magnitude (AM), acceleration duration (AD), significant acceleration frequency (SAF), turn magnitude (TM), turn duration (TD), and significant turn frequency (STF). A description of each measure is shown in Table 1.

**Table 1:**
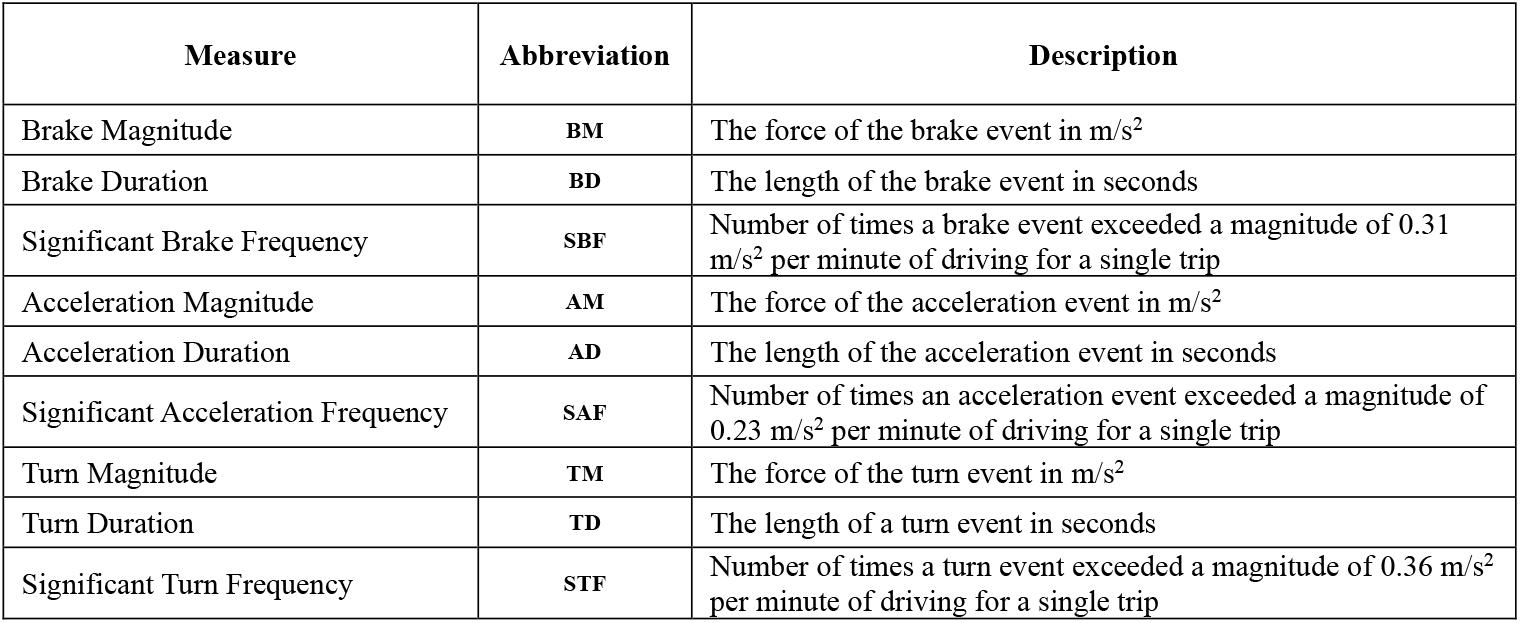
Driving Performance Measures.

| Measure | Abbreviation | Description |
| --- | --- | --- |
| Brake Magnitude | **BM** | The force of the brake event in $\text{m/s}^2$ |
| Brake Duration | **BD** | The length of the brake event in seconds |
| Significant Brake Frequency | **SBF** | Number of times a brake event exceeded a magnitude of $0.31 \text{ m/s}^2$ per minute of driving for a single trip |
| Acceleration Magnitude | **AM** | The force of the acceleration event in $\text{m/s}^2$ |
| Acceleration Duration | **AD** | The length of the acceleration event in seconds |
| Significant Acceleration Frequency | **SAF** | Number of times an acceleration event exceeded a magnitude of $0.23 \text{ m/s}^2$ per minute of driving for a single trip |
| Turn Magnitude | **TM** | The force of the turn event in $\text{m/s}^2$ |
| Turn Duration | **TD** | The length of a turn event in seconds |
| Significant Turn Frequency | **STF** | Number of times a turn event exceeded a magnitude of $0.36 \text{ m/s}^2$ per minute of driving for a single trip |

### Data retention and statistical analysis

Data were collected for 11,945 journeys completed by 97 participants. Three participants who were unable to produce usable driving performance data, and three participants who did not attend their OSA diagnostic appointment were excluded. On a pragmatic basis, trips less than 5 minutes (27% of the data) were excluded as likely too short to add useful event data. Additionally, journeys over 45 minutes (accounting for 8.5% of the data) were contributed disproportionately by a small number of participants and so were also excluded as unrepresentative. The final dataset was 7,834 journeys (over 2,000 hours of driving), from 91 participants. See Table 2. The mean number of trips per participant was 86, with a range of 3-716, see Figure 2.

**Table 2:** Data breakdown.

|  |  |
| --- | --- |
| n participants | 91 |
| Driving data |  |
| n trips | 7,834 |
| n brake events | 90,214 |
| n acceleration events | 199,388 |
| n turn events | 218,942 |
| n total events | 508,544 |
| Total driving time (hrs) | 2,042 |
| Mean trip duration (mins) | 15.6 |
| Diagnostic data |  |
| Mean ODI/AHI (SD / range) | 15 (15.8 / 0.1-84.2) |
| Mean ESS (SD / range) | 12.5 (5.3 / 0-24) |
| Demographics |  |
| Mean age years (SD / range) | 50.8 (13.5 / 22-77) |
| Sex F/M (% Female) | 32/59 (35.2%) |
| Mean BMI kg/m<sup>2</sup> (SD range) | 32.8 (7.7 / 22-58) |

**Figure 2:**
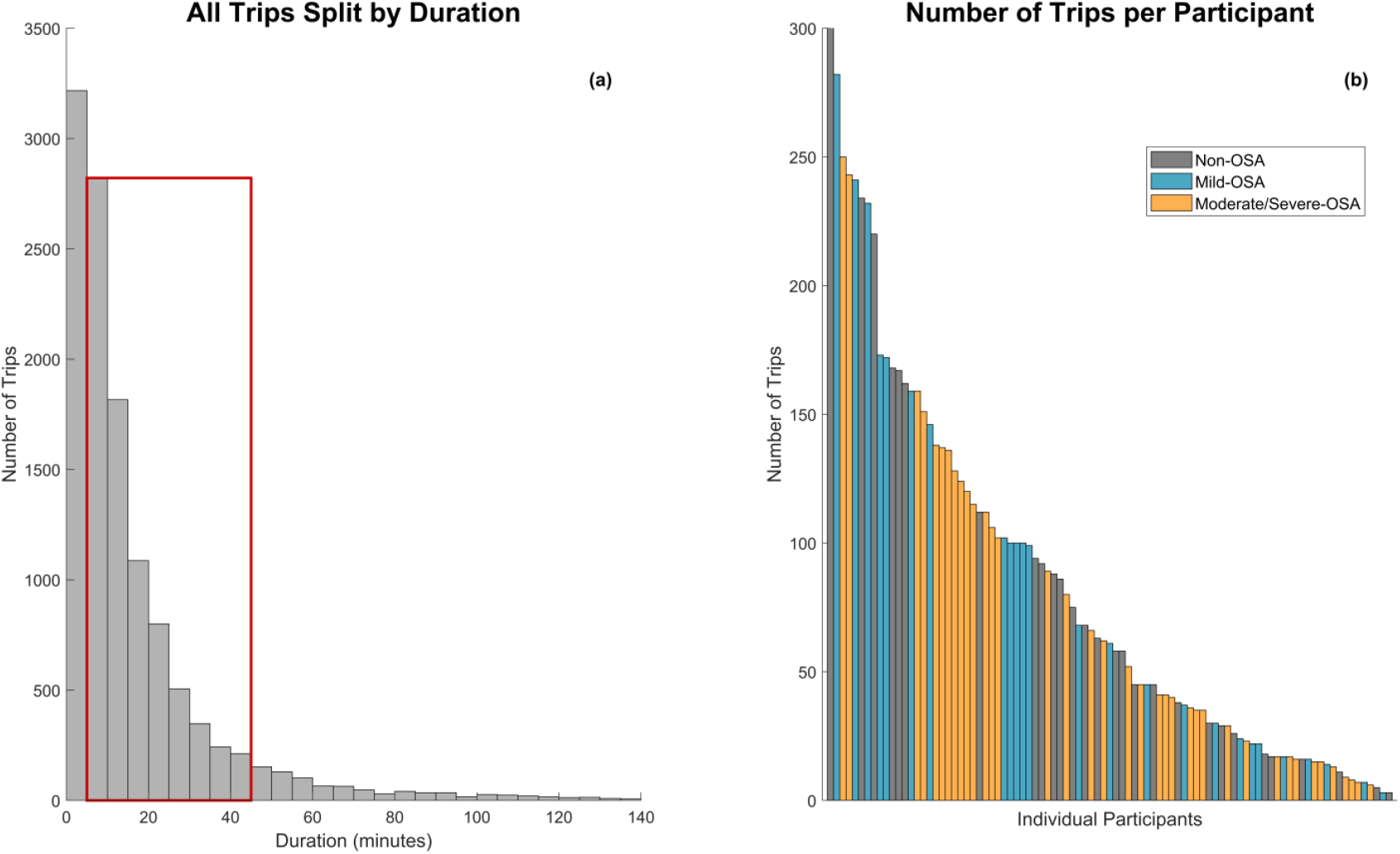
(a) Histogram showing the number of trips collected split by 5-minute intervals, cut at 140 minutes on the x-axis. The red box contains trips with a duration between 5-45 minutes, used for analysis. (b) Bar chart showing the ranked number of trips completed per participant, cut at 300 on the y-axis. Colour reflects the patient’s OSA severity. Non-OSA (ODI < 5): grey, Mild-OSA (ODI 5-15): blue, Moderate-Severe-OSA (ODI > 15): yellow.

The data were analysed using linear mixed-effects (LME) models including fixed effects: time into trip (not for frequency measures), trip duration, ODI, ESS, age, sex, BMI. Individuals were treated as random effects, controlling for different numbers of trips undertaken and other potential confounding factors across participants. Each analysis contained six or nine models (one for each driving measure, except when time into trip was measured and frequency measures could not be used). Effect size was calculated as the LME fixed effect estimate divided by the residual SD. For independent effects, continuous variables effect sizes were scaled by two standard deviations, to aid interpretability across the distribution, and comparison with categorical effect sizes, in line with recommendations (30). All p-values were Bonferroni-adjusted across nine driving measures for all models, including frequency measures. All analyses were conducted in MATLAB (version R2024a).

## Results

Table 2 contains a summary of the data retained for analysis. The first analysis tested which clinical markers of OSA and demographic factors relate to real-world driving peri-diagnosis. Only age had a significant effect on driving, but even here effect sizes were small, with lower acceleration magnitude (p < 0.05, effect size: -0.15), longer acceleration duration (p < 0.01, effect size: 0.16) and lower turn magnitude (p < 0.05, effect size: -0.16). ODI, ESS, sex and BMI were not found to be significant factors for any of our nine driving measures. Trip duration was a significant factor for all nine driving measures, and time into trip was significant for all 6 relevant measures. [See Figure 3 and supplementary Table a.] To quantify how much variance was due to stable individual differences versus within-person fluctuations, we computed the Intraclass Correlation Coefficient (ICC) from the LME’s random-intercept and residual standard deviations. Between-participant effects explained only 3-6% of the variance in magnitude and duration measures. But they explained 23-27% of the variance in the frequency measures. The within-participant variance may indicate day-to-day differences in performance but will be heavily influenced by different itineraries and traffic conditions on different days.

**Figure 3:**
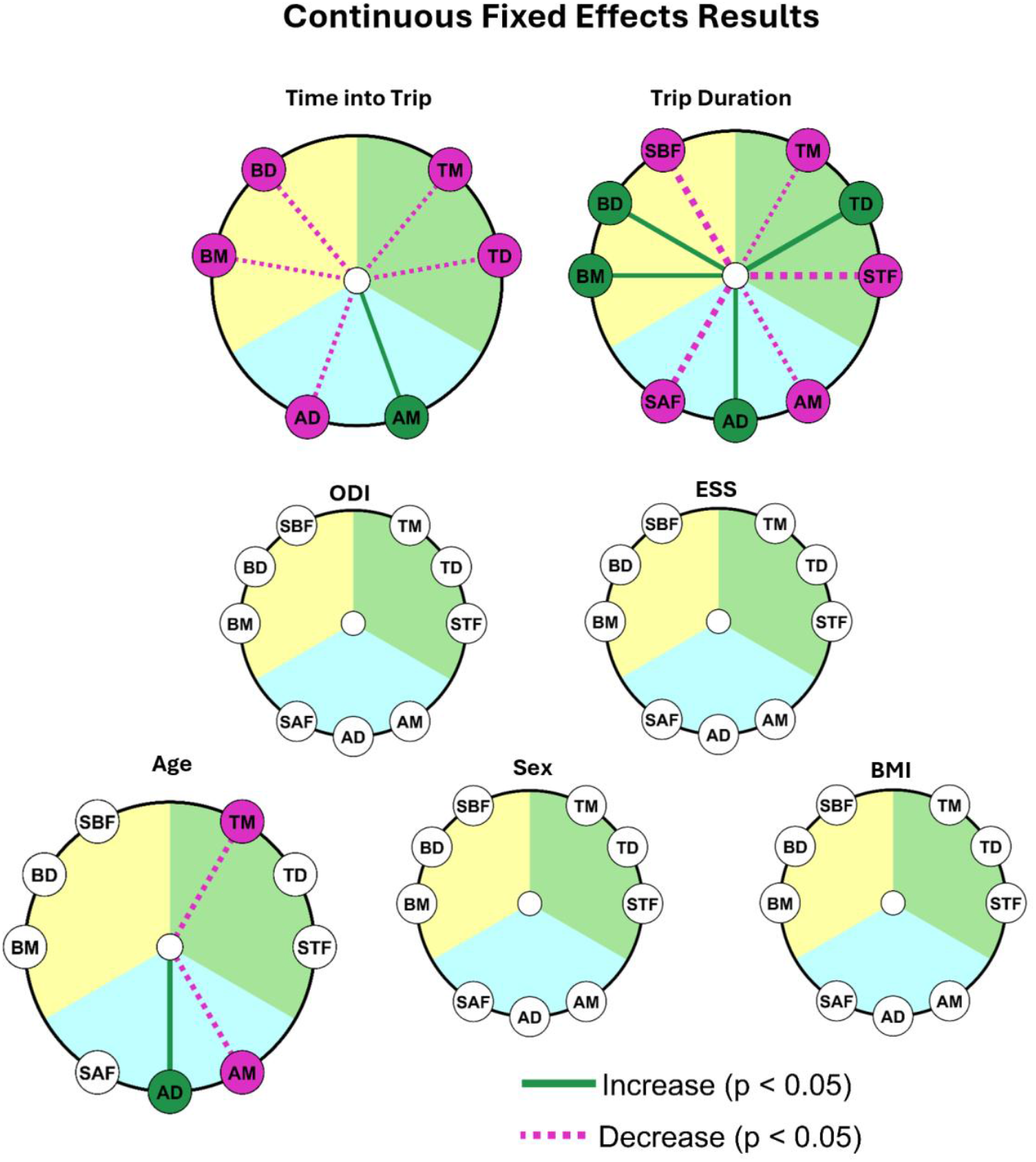
Radial significance plots of linear mixed-effects (LME) model results from peri-diagnosis testing. Each circle reflects independent fixed effects within models. Outer markers reflect driving measures tested, grouped by type: (Braking (yellow): BM = brake magnitude, BD = brake duration, SBF = significant brake frequency, Acceleration (blue): AM = acceleration magnitude, AD = acceleration duration, SAF = significant acceleration frequency and Turning (green): TM = turn magnitude, TD = turn duration, STF = significant turn frequency). Presence of a line indicates significance (p<0.05), as determined by the LME results after adjusting for multiple comparisons, green (increase), purple (decrease), width indicates effect size, cut at a minimum of 0.05 and maximum of 1.2.

In the second analysis participants were stratified based on categorical combinations of OSA severity and sleepiness. OSA was categorised as absent (ODI <5), mild (ODI ≥5 and < 15) and moderate-severe (ODI ≥ 15). Sleepiness was deemed as absent (ESS <11) or present (ESS ≥11). Participants without OSA and not sleepy were taken as the comparator (best control). Turn magnitude was reliably higher than the comparator for all groups: non-OSA / EDS (p < 0.01, effect size: 0.33), mild-OSA / non-EDS (p < 0.01, effect size: 0.34), mild-OSA / EDS (p < 0.05, effect size: 0.32), moderate-severe-OSA / non-EDS (p < 0.01, effect size: 0.35), moderate-severe-OSA / EDS (p < 0.01, effect size: 0.37). Significant turn frequency was reliably higher than the comparator for: non-OSA/ EDS (p < 0.05, effect size: 0.68), mild-OSA / non-EDS (p < 0.05, effect size: 0.66), moderate-severe-OSA / non-EDS (p < 0.05, effect size: 0.74), moderate-severe-OSA / EDS (p < 0.01, effect size: 0.77). The mild-OSA / EDS group was not different from the comparator group, after multiple comparisons correction. [See Figure 4 and supplementary Table b.]

**Figure 4:**
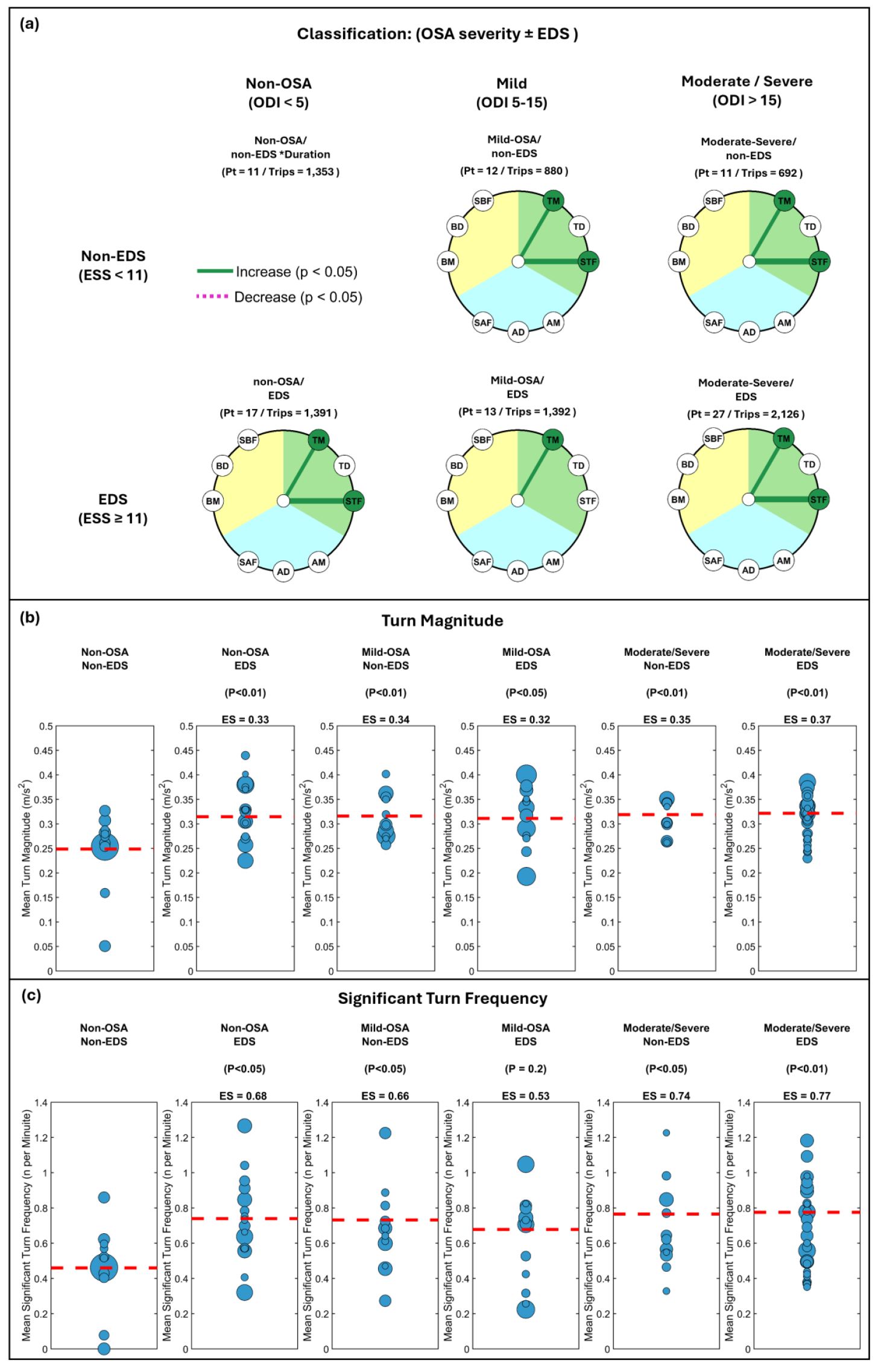
(a) Radial significance plots of linear mixed-effects (LME) model results from peri-diagnosis classification testing. Each circle reflects independent fixed effects within models (including each categorical classification, except non-OSA/non-sleepy which all other classifications were compared to). Outer markers reflect driving measures tested, grouped by type: (Braking (yellow): BM = brake magnitude, BD = brake duration, SBF = significant brake frequency, Acceleration (blue): AM = acceleration magnitude, AD = acceleration duration, SAF = significant acceleration frequency and Turning (green): TM = turn magnitude, TD = turn duration, STF = significant turn frequency). Presence of a line indicates significance (p<0.05), as determined by the LME results after adjusting for multiple comparisons, green (increase), purple (decrease), width indicates effect size. cut at a minimum of 0.05 and maximum of 1.2. (b) Participant-level mean turning magnitude, stratified by clinical classification. Marker size is proportional to the number of journeys per participant. The dashed red line shows the reference (mean of participant means in the non-OSA / non-EDS group); for other groups it shows the reference plus the group effect from the linear mixed-effects (LME) model. (c) Participant-level mean significant turn frequency, stratified by clinical classification. Marker size is proportional to the number of journeys per participant. The dashed red line shows the reference (mean of participant means in the non-OSA / non-EDS group); for other groups it shows the reference plus the group effect from the linear mixed-effects (LME) model

Because the number of participants in the Non-OSA/Non-EDS comparator group was small (n=11), yet contributed a large number of journeys (n=1,353), and anchors all contrasts, we ran a control-constrained leave-one-out (LOO) robustness check: models were re-fitted 11 times, each time omitting one comparator participant. For turn magnitude and significant turn frequency, 89 and 81% (respectively) of comparisons remained statistically significant, and under worst-case omissions, effect sizes remained moderate and clinically meaningful. [See Supplementary text a, and Supplementary Figures a and b.]

Furthermore, we ran additional analyses testing both the interaction effect of ODI and ESS, uncategorised, and stratification based on clinical outcome: CPAP starter, discharged from clinic, follow-up appointment without CPAP and treated with mandibular advancement device (MAD). Neither of these analyses showed a reliable effect for any of the nine driving measures. [See Supplementary Tables c and d.] Thus, stratifying patients by classifications using both standard OSA severity and EDS thresholds identified between-group differences in driving behaviour more effectively than either continuous ESS/ODI or outcome-based stratification. Consistent with this, the classification model had substantially better support on the AIC scale (Akaike weights of 0.989 vs 0.011 for the continuous model), supporting the categorical framework for patient classification. [See Supplementary text b.]

As time into trip and trip duration were significant predictors of all driving performance measures in our first analysis we ran two independent models testing interaction effects of both time measures with ODI and ESS. ODI and time into trip interactions were not reliably different for any measurement. ODI and duration interactions were significant for 3/9 measures but effect sizes were not clinically relevant (range: 0.02 - 0.03). For the ESS models, there were no significant interactions for ESS and time into trip. ESS and duration interactions were statistically significant for 4/9 measures, but effect sizes were again not clinically relevant (range: 0.03 - 0.05). [See Supplementary Figure c and supplementary Tables e and f.]

## Discussion

We analysed data collected from 7,834 real-world journeys (comprising in total more than half a million braking, acceleration and turning events) from 91 patients at peri-diagnosis for OSA, to test whether clinical markers of OSA (ODI and ESS) and demographics were related to driving behaviour. In our first model, only age, time into trip and trip duration were associated with changes in our driving measures, while continuous ODI and ESS were not differentiators of driving behaviours. In our second model, when stratifying our participants into categories by OSA severity and presence or absence of EDS, we showed that for all groups, turning was more forceful compared to the non-OSA / non-EDS group.

Our first analysis showed that older age is associated with reduced acceleration and turn magnitude, and longer acceleration duration. Within our sample age range (22-77 years), this pattern suggests younger drivers had a more dynamic and perhaps riskier driving style. Previous studies have shown that there are complex interactions between age and driving exposure including distances driven and the amount of night driving but by and large, and perhaps convergently, drivers under the age of 30 are more likely to be involved in crashes (31).

We observed substantial within-participant variance for driving metrics, plausibly reflecting varying itineraries (urban and motorway driving proportions) and day-to-day traffic density beyond driving styles. While our first model showed that time into trip and trip duration were each related to all our tested driving measures, the effect sizes were small. ‘Time on task’ may explain time-into-trip effects, whereas the duration effect likely reflects varying road conditions, with long and short journeys containing different urban–motorway proportions.

Critically, for legal and clinical decision making, categorising participants by OSA severity and presence or absence of sleepiness, revealed that relative to the non-OSA / non-EDS group, all other groups displayed a higher level of turn magnitude. Moreover, all groups except the mild-OSA / EDS group, also had a higher rate of significant turn frequency. These findings remained robust in LOO checks, which indicated that clinically relevant effect sizes were preserved, even under worst-case participant omissions.

These results partially converge with prior work relating OSA severity and subjective sleepiness to driving outcomes. Studies that have tested OSA severity and sleepiness separately have produced inconsistent results (1), and our analysis suggests using a combined classification framework could reduce confounding factors that may arise when testing OSA severity or sleepiness in isolation. This is pertinent, because patients present to clinic with all combinations of OSA severity and daytime sleepiness (32). Notably, our data also indicated that the non-OSA / EDS group differed from the non-OSA/non-EDS group, suggesting that clinic attendees without OSA who report high sleepiness may exhibit impairments comparable to those with OSA, with or without EDS, calling for a revision of the fit-to-drive criteria.

A limitation of our classification analysis was the small number of patients with severe OSA without EDS, which forced us to group moderate and severe cases together to maintain meaningful sized groups, which may have reduced power to detect effects unique to severe OSA with and without EDS. In addition, applying standard clinical thresholds yielded uneven group sizes (e.g., moderate-severe / EDS n = 27 vs. moderate-severe / non-EDS n = 11), contributing to unequal numbers of trips (2,126 and 692, respectively). Our mixed-effects models should have mitigated these imbalances in statistical testing, but residual bias cannot be excluded.

Across our clinical classification analysis, turning rather than braking or accelerating behaviour was consistently higher in magnitude among clinical groups relative to the non-OSA / non-EDS group. Turning places heavy demands on gaze-steering coupling and preview control, including the use of tangent point and future path point information (33), processes that are less central to longitudinal control (straight-line control). Although causality cannot be inferred, this selective difference could reflect that turning-related cognition may be more affected by OSA and EDS than other aspects of driving. This aligns with evidence showing that cognitive processes degrade at different rates during sleep onset (34), and that novice drivers exhibit narrower horizontal visual search than experienced drivers (35). Although turn manoeuvres are well known to be crash-prone, there is little work validating turning-specific telematics as predictors of crash risk. Our findings indicate turning behaviour differentiates clinical groups and future studies should test these metrics against crashes and near-misses.

From our data we cannot determine whether underlying conditions influenced which journeys drivers elected to undertake. In addition, some drivers may have adopted compensatory strategies that mask impairment signals from the driving data. In a simulator study, which used an AHI > 10 to demarcate clinically relevant OSA, patients were found to be either “vulnerable” or “resistant” to performance decline under extended wakefulness (9). It is unclear whether individuals with apparently normalised driving profiles are genuinely unimpaired, or if differences in susceptibility to sleepiness makes commonly used compensatory strategies such as opening the window or drinking coffee (36) more or less effective. Future work should test whether drivers with severe OSA and EDS, who appear normalised have cognitive compensation and/or whether coping strategies play a role in risk reduction.

Given the broad associations of time into trip and trip duration with our performance measures, we examined interactions between time measures and clinical markers. The interaction between trip duration and ODI was reliably associated with 3/9 measures, and the interaction between trip duration and ESS with 4/9 measures. However, estimated effects were minimal and unlikely to be clinically meaningful. The directions of change differed (e.g., increasing trip duration with higher ODI was linked to decreased turning magnitude, whereas increasing trip duration with higher ESS was linked to increased turning magnitude) raising the possibility that OSA severity and EDS affect driving via distinct mechanisms. This interpretation is tentative and requires replication. It may also explain why no effect emerged when assessing the direct ODI and ESS interactions. Incorporating journey length and context may help reveal such nuanced effects.

A key strength of our study is its real-world setting. Unlike crash report analyses, which capture only rare, catastrophic events, or simulator experiments, which despite high data density, lack real-world validity (22), our telematics approach recorded behaviour across entire trips at scale (over half a million braking, acceleration, and turning events). This demonstrates that the methodology used can characterise situationally dependent impairment in real-world driving. However, at the same time, real-world complexity complicates risk inference, as the same metrics (e.g., fast acceleration or late braking) can be adaptive in one context and unsafe in another. Consequently, while challenging, progressing beyond uninformative laboratory-based assessments towards an ecologically valid measurement of true real-world behaviour is essential.

These findings carry important implications for clinical and regulatory practice. In the UK, drivers with confirmed moderate or severe OSA accompanied by excessive sleepiness must cease driving and notify the DVLA, and may only resume once their condition is controlled and treatment adhered to, with periodic medical review required.

Similarly, under EU Directive 2014/85/EU implemented in 2015, individuals with suspected moderate-to-severe OSA must undergo further medical evaluation before obtaining or renewing a licence, and may only be permitted to drive if they demonstrate effective treatment adherence and improvement in sleepiness, along with regular re-evaluation. Although ten years on from inception, implementation of these regulations has been problematic and not consistently applied across EU member countries (37).

Our results of turning behaviour metrics that can distinguish high-risk drivers (both OSA and non-OSA but sleepy individuals) suggest a shift from current regulations which focus on diagnosis and self-report toward objective behavioural indicators of impairment. This provides a promising path to refine licensing decisions, while avoiding unnecessary driving restrictions. For example, telematics could help identify safely treated patients, and capture improvements post-therapy that traditional assessment measures may overlook.

Future research should identify combinations of telematics metrics that best predict crash and near-miss risk (including what additional driving contextual data assist model accuracy), and quantify how much data per participant is required to deliver personalised risk estimates. The apparent elevation in risk among clinic attendees without OSA, but with high EDS also warrants targeted studies. Beyond diagnosis, telematics may also be suitable for tracking functional improvements after therapy initiation or to flag deterioration that self-report misses. Prospective designs that link telematics to verified outcomes (e.g., insurance claims) and explicitly model journey context (road class, traffic density, time of day, weather) and driving exposure will be important for developing personalised risk monitoring in OSA and general sleepiness.

## Data Availability

De-identified aggregated driving metrics and analysis code may be made available on request to qualified researchers for non-commercial use under a data-use policy, from publication until 36 months thereafter (contact). Individual-level raw telematics, demographic and medical data will not be shared due to privacy constraints.

## Contributors

KL, TA, and IS conceptualised the study, KL coordinated and delivered study implementation, completed analysis, and wrote the first draft of the manuscript. TA, and IS provided supervision of the study set-up, delivery and implementation, reviewed and revised earlier versions of the manuscript, and provided expert clinical and analysis advice. IS and TA secured funding to undertake the study.

All authors had full access to and verified the underlying data in the study, full access to the aggregated results and approved the decision to submit.

## Acknowledgements

We would like to express appreciation to all the participants who volunteered to take part in the study, Royal Papworth Charity for financial support, and all departments across the University of Cambridge and Royal Papworth Hospital for facilitating study set-up and delivery.

## References

1. Ellen R, Marshall S, Palayew M, Molnar F, Wilson K, Man-Son-Hing M. Systematic review of motor vehicle crash risk in persons with sleep apnea. J Clin Sleep Med. 2006;2(2):193–200.

2. Tregear S, Reston J, Schoelles K, Phillips B. Obstructive sleep apnea and risk of motor vehicle crash: systematic review and meta-analysis. J Clin Sleep Med. 2009 Dec 15;5(6):573–81.

3. Barbé F, Sunyer J, de la Peña A, Pericas J, Mayoralas LR, Antó JM, et al. Effect of continuous positive airway pressure on the risk of road accidents in sleep apnea patients. Respiration. 2007;74(1):44–9.

4. Masa JF, Rubio M, Findley LJ. Habitually sleepy drivers have a high frequency of automobile crashes associated with respiratory disorders during sleep. Am J Respir Crit Care Med. 2000 Oct;162(4 Pt 1):1407–12.

5. Mulgrew AT, Nasvadi G, Butt A, Cheema R, Fox N, Fleetham JA, et al. Risk and severity of motor vehicle crashes in patients with obstructive sleep apnoea/hypopnoea. Thorax. 2008 June;63(6):536–41.

6. Terán-Santos J, Jiménez-Gómez A, Cordero-Guevara J. The association between sleep apnea and the risk of traffic accidents. Cooperative Group Burgos-Santander. N Engl J Med. 1999 Mar 18;340(11):847–51.

7. Lloberes P, Levy G, Descals C, Sampol G, Roca A, Sagales T, et al. Self-reported sleepiness while driving as a risk factor for traffic accidents in patients with obstructive sleep apnoea syndrome and in non-apnoeic snorers. Respir Med. 2000 Oct;94(10):971–6.

8. Shiomi T, Arita AT, Sasanabe R, Banno K, Yamakawa H, Hasegawa R, et al. Falling asleep while driving and automobile accidents among patients with obstructive sleep apnea-hypopnea syndrome. Psychiatry Clin Neurosci. 2002 June 1;56(3):333–4.

9. Vakulin A, Green MA, D’Rozario AL, Stevens D, Openshaw H, Bartlett D, et al. Brain mitochondrial dysfunction and driving simulator performance in untreated obstructive sleep apnea. J Sleep Res [Internet]. 2022 Apr 1;31(2). Available from: 10.1111/jsr.13482

10. Pizza F, Contardi S, Mondini S, Trentin L, Cirignotta F. Daytime Sleepiness and Driving Performance in Patients with Obstructive Sleep Apnea: Comparison of the MSLT, the MWT, and a Simulated Driving Task. Sleep. 2009;32(3):382–91.

11. Engleman HM, Asgari-Jirhandeh N, McLeod AL, Ramsay CF, Deary IJ, Douglas NJ. Self-reported use of CPAP and benefits of CPAP therapy: a patient survey. Chest. 1996 June;109(6):1470–6.

12. Findley L, Unverzagt M, Guchu R, Fabrizio M, Buckner J, Suratt P. Vigilance and automobile accidents in patients with sleep apnea or narcolepsy. Chest. 1995 Sept;108(3):619–24.

13. Noda A, Yagi T, Yokota M, Kayukawa Y, Ohta T, Okada T. Daytime sleepiness and automobile accidents in patients with obstructive sleep apnea syndrome. Psychiatry Clin Neurosci. 1998 Apr 1;52(2):221–2.

14. Coelho J, Bailly S, Baillieul S, Sagaspe P, McNicholas WT, Taillard J, et al. Predictors of driving risk in patients with obstructive sleep apnea syndrome treated by continuous positive airway pressure: a French multicenter prospective cohort. SLEEP [Internet]. 2024 Nov 8;47(11). Available from: https://academic.oup.com/sleep/article/doi/10.1093/sleep/zsae211/7774885

15. Barbé, Pericás J, Muñoz A, Findley L, Antó JM, Agustí AG. Automobile accidents in patients with sleep apnea syndrome. An epidemiological and mechanistic study: An epidemiological and mechanistic study. Am J Respir Crit Care Med. 1998 July;158(1):18–22.

16. Flemons WW, Remmers JE, Whitelaw WA. The correlation of a computer simulated driving program with polysomnographic indices and neuropsychological tests in consecutively referred patients for assessment of sleep apnea. Sleep. 1993 Dec;16(8 Suppl):S71.

17. Rizzo D, Baltzan M, Grad R, Postuma R. Risk of OSA affects reaction time and driving performance more than insomnia in the Canadian Longitudinal Study on Aging. Transp Res Part F Traffic Psychol Behav. 2023 May 1;95:261–70.

18. Turkington PM, Sircar M, Allgar V, Elliott MW. Relationship between obstructive sleep apnoea, driving simulator performance, and risk of road traffic accidents. Thorax. 2001;56:800–5.

19. George CF, Boudreau AC, Smiley A. Simulated driving performance in patients with obstructive sleep apnea. Am J Respir Crit Care Med. 1996 July;154(1):175–81.

20. Ghosh D, Jamson SL, Baxter PD, Elliott MW. Continuous measures of driving performance on an advanced office-based driving simulator can be used to predict simulator task failure in patients with obstructive sleep apnoea syndrome. Thorax. 2012 Sept;67(9):815–21.

21. Pichel F, Zamarrón C, Magán F, Rodríguez JR. Sustained attention measurements in obstructive sleep apnea and risk of traffic accidents. Respir Med. 2006 June;100(6):1020–7.

22. Bonsignore MR, Randerath W, Schiza S, Verbraecken J, Elliott MW, Riha R, et al. European Respiratory Society statement on sleep apnoea, sleepiness and driving risk [Internet]. Vol. 57, European Respiratory Journal. European Respiratory Society; 2021. Available from: 10.1183/13993003.01272-2020

23. Drowsy Driving: A National Sleep Foundation Position Statement and Call to Action [Internet]. National Sleep Foundation. 2023 [cited 2025 Sept 11]. Available from: https://www.thensf.org/wp-content/uploads/2023/11/NSF-Position-Statement_Drowsy-Driving_11.06.23.pdf

24. The Lancet Respiratory Medicine. Driving forwards in the management of OSA. Lancet Respir Med. 2025 Sept;13(9):769.

25. The Lancet Respiratory Medicine. Sleep apnoea and driving: more clarity needed. Lancet Respir Med. 2017 Feb 1;5(2):85.

26. Baecke P, Bocca L. The value of vehicle telematics data in insurance risk selection processes. Decis Support Syst. 2017 June 1;98:69–79.

27. Chandar B, Gneezy U, List JA, Muir I. The Drivers of Social Preferences: Evidence from a Nationwide Tipping Field Experiment [Internet]. 2019. Available from: http://www.nber.org/papers/w26380

28. Babulal GM, Johnson A, Fagan AM, Morris JC, Roe CM. Identifying preclinical Alzheimer’s disease using everyday driving behavior: Proof of concept. J Alzheimers Dis. 2021;79(3):1009– 14.

29. Doherty JM, Roe CM, Murphy SA, Johnson AM, Fleischer E, Toedebusch CD, et al. Adverse driving behaviors are associated with sleep apnea severity and age in cognitively normal older adults at risk for Alzheimer’s disease. Sleep [Internet]. 2022 June 1;45(6). Available from: 10.1093/sleep/zsac070

30. Gelman A. Scaling regression inputs by dividing by two standard deviations. Stat Med. 2008 July 10;27(15):2865–73.

31. Regev S, Rolison JJ, Moutari S. Crash risk by driver age, gender, and time of day using a new exposure methodology. J Safety Res. 2018 Sept 1;66:131–40.

32. Ye L, Pien GW, Ratcliffe SJ, Björnsdottir E, Arnardottir ES, Pack AI, et al. The different clinical faces of obstructive sleep apnoea: A cluster analysis. European Respiratory Journal. 2014 Dec 1;44(6):1600–7.

33. Okafuji Y, Fukao T. Theoretical interpretation of drivers’ gaze strategy influenced by optical flow. Sci Rep. 2021 Jan 27;11(1):2389.

34. Lacaux C, Strauss M, Bekinschtein TA, Oudiette D. Embracing sleep-onset complexity. Trends Neurosci. 2024 Apr 1;47(4):273–88.

35. Robbins C, Chapman P. How does drivers’ visual search change as a function of experience? A systematic review and meta-analysis. Accid Anal Prev. 2019 Nov;132:105266.

36. Dwarakanath A, Palissery V, Ghosh D, Jamson S, Elliott M. An exploratory study evaluating the use of coping strategies while driving in obstructive sleep apnoea syndrome patients and controls. ERJ Open Res [Internet]. 2024 Jan 22 [cited 2025 June 4];10(1). Available from: 10.1183/23120541.00638-2023

37. McNicholas WT, van der Werf YD, Hartley S, Philip P, study collaborators in the Assembly of National Sleep Societies and other National Representatives. Implementation of European national driving regulations for obstructive sleep apnoea: challenges and recommendations. Eur Respir J. 2025 July 24;66(1):2402484.

