## Supplementary Material for "A Turn in the Road: Real-life Driving Differences in Obstructive Sleep Apnoea and Excessive Daytime Sleepiness"

### 1 Supplementary Text

#### 1.1  Supplementary Text a: Leave One Out stress testing of Clinical Classification Results

For turn magnitude we evaluated 55 LOO refits (5 significant group contrasts × 11 omissions), with 89% (49/55) comparisons remaining statistically significant (using the same criteria as the main analysis), and the direction of effect was consistent across all iterations. Excluding one influential comparator participant decreased statistical significance across all contrasts, however the corresponding effect sizes under this most stringent test were still clinically meaningful (range: 0.22 – 0.26). For significant turn frequency we evaluated 44 LOO refits (4 significant group contrasts × 11 omissions), with 81% (36/44) of comparisons remaining significant, with effect directions consistent across all iterations. Two comparator participants had a modest impact (the mild-OSA / non-EDS contrast became non-significant), and two others had a larger influence, after which only the moderate-severe / EDS contrast retained significance. Even in the worst-case omission, effects for the groups showing main effects remained moderate and clinically meaningful (range: 0.56 – 0.65).

#### 1.2  Supplementary Text b: Model fit comparison: Continuous vs Classification

Model comparison favoured the categorical classification over the continuous-interaction specification: AIC −87,798 vs −87,789 (ΔAIC = 9). The Akaike Information Criterion (AIC = 2k − 2·log-likelihood) balances fit and parsimony; smaller is better, and only differences are interpretable. On the AIC scale, a 9-unit gap constitutes strong evidence for the lower-AIC model: the relative likelihood of the alternative is exp (−ΔAIC/2) = exp (−4.5) ≈ 0.011, yielding Akaike weights 0.989 vs 0.011 (≈90:1 support). Because AIC already penalises model complexity, the absolute magnitude (or sign) of AIC values is immaterial; what matters is the ΔAIC. These findings support a categorical framework for patient classification.

### 2 Supplementary Figures

#### 2.1  Supplementary Figure a: Leave One Out results (Turn Magnitude)

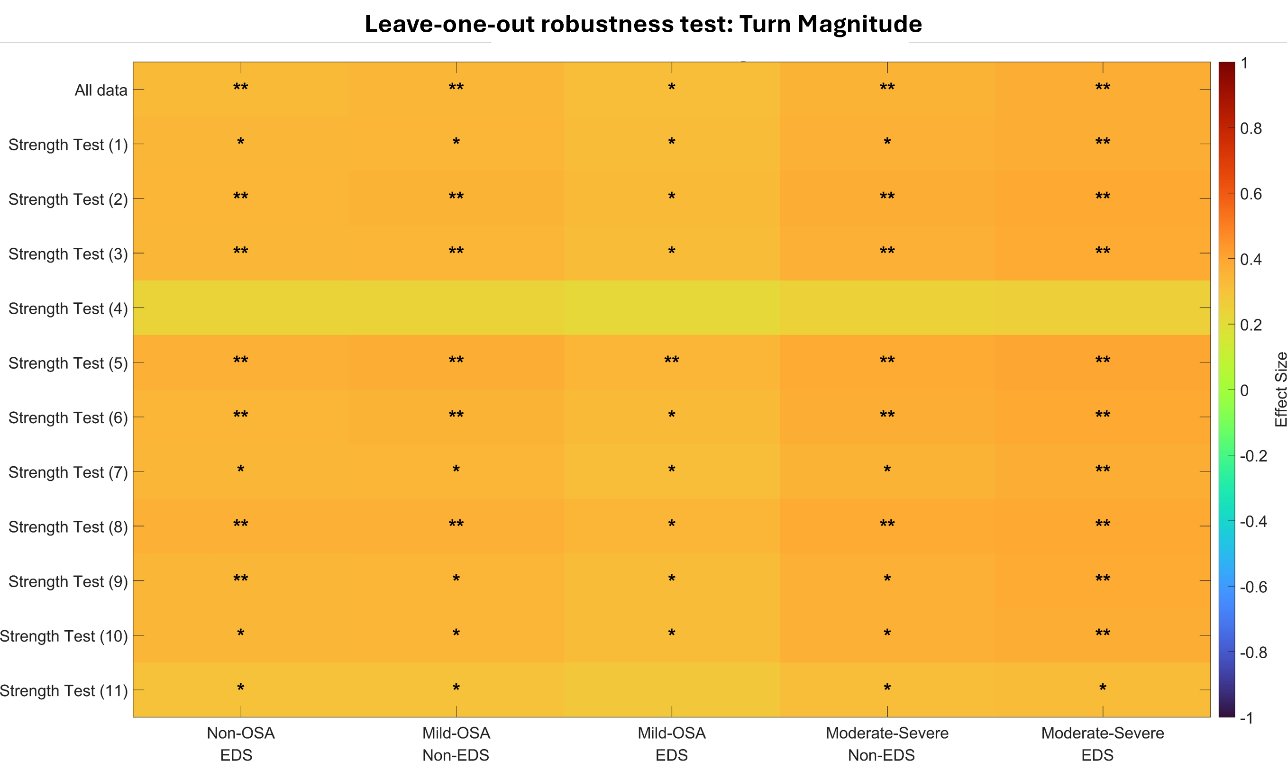

Supplementary Figure a. Heatmap of leave-one-out (LOO) robustness for clinical-classification effects on turning magnitude. Rows show the full model and models with each participant omitted in turn. Columns are clinical categories contrasted with the non-OSA/non-EDS reference. Asterisks indicate significance (* p < 0.05; ** p < 0.01).

#### 2.2 Supplementary Figure b: Leave-One-Out results (Significant Turn Frequency)

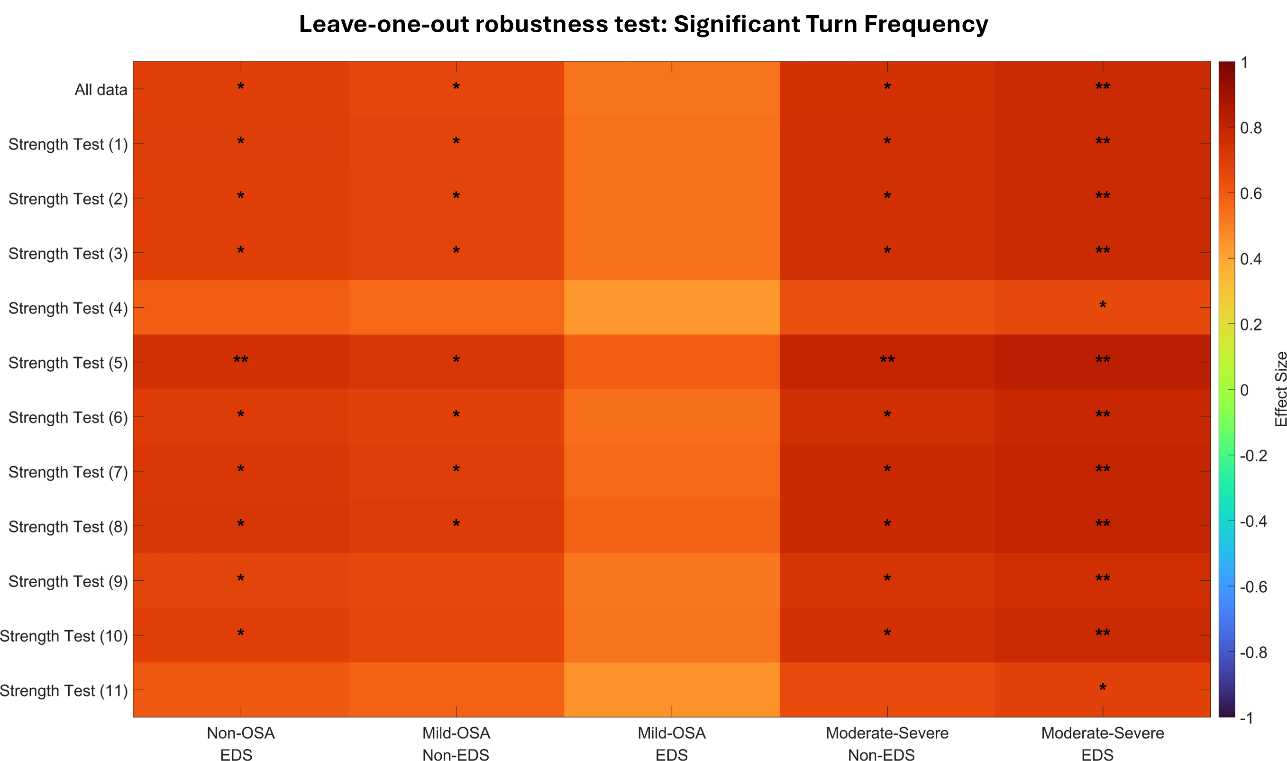

Supplementary Figure b. Heatmap of leave-one-out (LOO) robustness for clinical-classification effects on significant turn frequency. Rows show the full model and models with each participant omitted in turn. Columns are clinical categories contrasted with the non-OSA/non-EDS reference. Asterisks indicate significance (* p < 0.05; ** p < 0.01).

#### 2.3  Supplementary Figure c: ODI and ESS Interactions with time variables

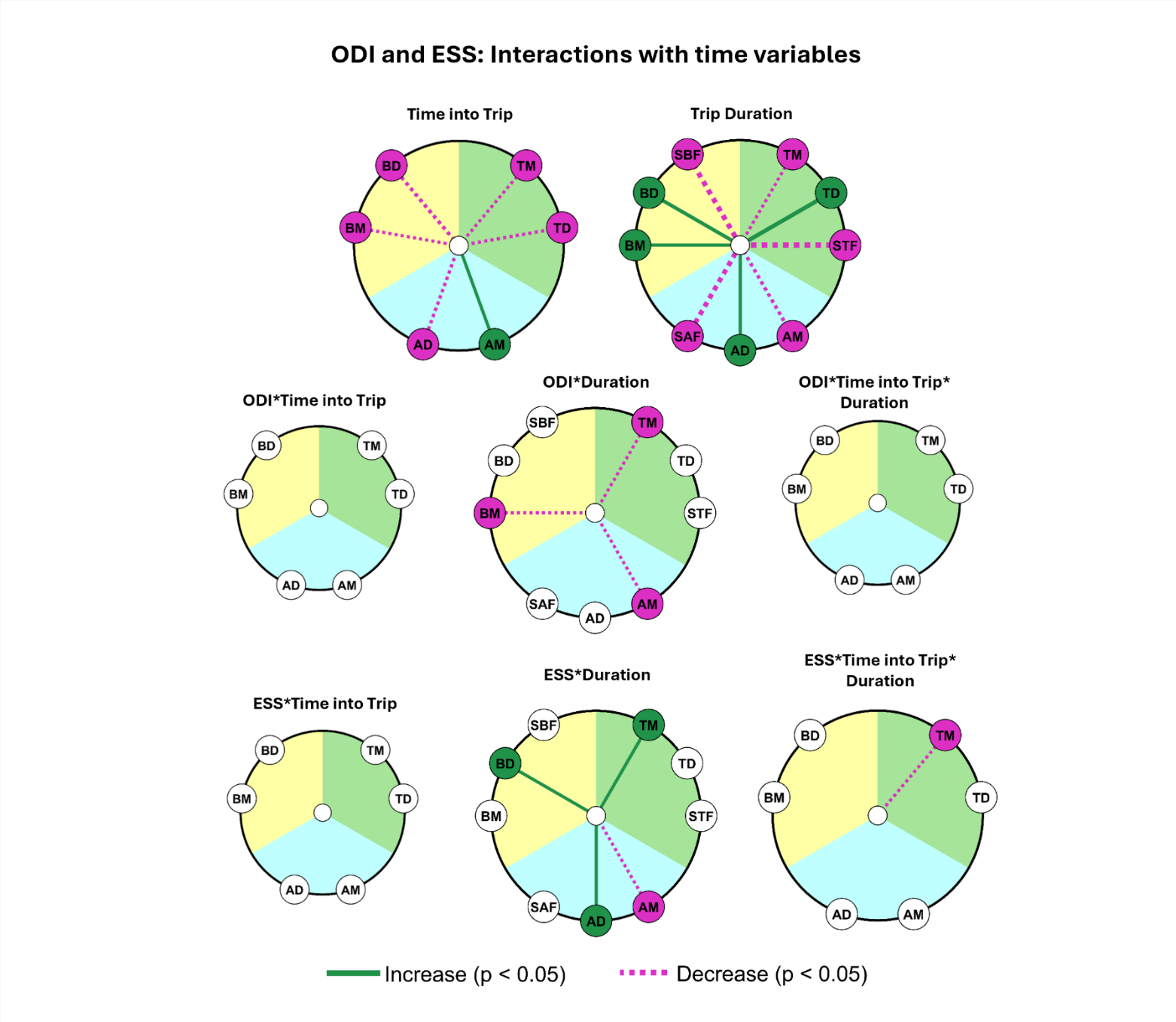

Supplementary Figure c: Radial significance plots of linear mixed-effect (LME) model results. Top row reflects time into trip and trip duration results from non-interaction analyses (also displayed in Figure 3). Middle (ODI), and bottom (ESS), rows reflect OSA clinical marker interactions with time variables. Each circle reflects independent interaction effects within models. Outer markers reflect driving measures tested, grouped by type: (Braking (yellow): BM = brake magnitude, BD = brake duration, SBF = significant brake frequency, Acceleration (blue): AM = acceleration magnitude, AD = acceleration duration, SAF = significant acceleration frequency and Turning (green): TM = turn magnitude, TD = turn duration, STF = significant turn frequency). Presence of a line indicates significance, green (increase), purple (decrease), width indicates effect size. cut at a minimum of 0.05 and maximum of 1.2.

### 3 Supplementary Tables

#### 3.1 Supplementary Table a: Continuous Fixed Effects Factor LME Results

| **Event** | **Feature** | **Predictor** | **Estimate** | **DF** | **P** | **Adjusted**  **P** | **Effect**  **Size (LME)** | **Data Points** | **Pt**  **(n)** |
| --- | --- | --- | --- | --- | --- | --- | --- | --- | --- |
| Brake | Magnitude | Intercept | 0.3964 | 90206 | 4.68E-85 | 4.21E-84 | 3.14 | 90214 | 91 |
|  |  | Time into Trip | -0.0003 | 90206 | 2.65E-08 | 2.38E-07 | -0.05 | 90214 | 91 |
|  |  | Trip Duration | 0.0002 | 90206 | 0.002 | 0.019 | 0.03 | 90214 | 91 |
|  |  | ODI | -0.0002 | 90206 | 0.395 | 1 | -0.04 | 91 | 91 |
|  |  | ESS | -0.0005 | 90206 | 0.344 | 1 | -0.04 | 91 | 91 |
|  |  | Age (Yrs) | -0.0006 | 90206 | 0.007 | 0.061 | -0.12 | 91 | 91 |
|  |  | Sex (F) | 0.0026 | 90206 | 0.683 | 1 | 0.02 | 91 | 91 |
|  |  | BMI (kg/m^2^) | -0.0004 | 90206 | 0.342 | 1 | -0.05 | 91 | 91 |
|  | Duration | Intercept | 5.2943 | 90206 | 1.28E-25 | 1.16E-24 | 1.60 | 90214 | 91 |
|  |  | Time into Trip | -0.0209 | 90206 | 4.58E-38 | 4.12E-37 | -0.11 | 90214 | 91 |
|  |  | Trip Duration | 0.0162 | 90206 | 3.92E-30 | 3.53E-29 | 0.10 | 90214 | 91 |
|  |  | ODI | 0.0007 | 90206 | 0.884 | 1 | 0.01 | 91 | 91 |
|  |  | ESS | 0.0168 | 90206 | 0.216 | 1 | 0.05 | 91 | 91 |
|  |  | Age (Yrs) | 0.0144 | 90206 | 0.010 | 0.092 | 0.10 | 91 | 91 |
|  |  | Sex (F) | 0.1513 | 90206 | 0.333 | 1 | 0.05 | 91 | 91 |
|  |  | BMI (kg/m^2^) | 0.0081 | 90206 | 0.451 | 1 | 0.03 | 91 | 91 |
|  | Sig Frequency | Intercept | 0.7125 | 7825 | 9.92E-07 | 8.93E-06 | 2.45 | 7832 | 91 |
|  |  | Trip Duration | -0.0042 | 7825 | 8.37E-30 | 7.53E-29 | -0.27 | 7832 | 91 |
|  |  | ODI | 0.0004 | 7825 | 0.776 | 1 | 0.04 | 91 | 91 |
|  |  | ESS | 0.0022 | 7825 | 0.565 | 1 | 0.08 | 91 | 91 |
|  |  | Age (Yrs) | -0.0020 | 7825 | 0.206 | 1 | -0.16 | 91 | 91 |
|  |  | Sex (F) | 0.0342 | 7825 | 0.446 | 1 | 0.12 | 91 | 91 |
|  |  | BMI (kg/m^2^) | -0.0049 | 7825 | 0.118 | 1 | -0.24 | 91 | 91 |
| Acceleration | Magnitude | Intercept | 0.2616 | 199380 | 3.79E-31 | 3.41E-30 | 2.23 | 199388 | 91 |
|  |  | Time into Trip | 0.0002 | 199380 | 6.52E-05 | 0.001 | 0.02 | 199388 | 91 |
|  |  | Trip Duration | -0.0006 | 199380 | 5.27E-66 | 4.75E-65 | -0.10 | 199388 | 91 |
|  |  | ODI | 7.95E-05 | 199380 | 0.708 | 1 | 0.02 | 91 | 91 |
|  |  | ESS | -8.86E-05 | 199380 | 0.882 | 1 | -0.01 | 91 | 91 |
|  |  | Age (Yrs) | -0.0008 | 199380 | 0.002 | 0.015 | -0.15 | 91 | 91 |
|  |  | Sex (F) | -0.0075 | 199380 | 0.274 | 1 | -0.06 | 91 | 91 |
|  |  | BMI (kg/m^2^) | 6.97E-05 | 199380 | 0.884 | 1 | 0.01 | 91 | 91 |
|  | Duration | Intercept | 3.6739 | 199380 | 7.57E-12 | 6.82E-11 | 1.25 | 199388 | 91 |
|  |  | Time into Trip | -0.0130 | 199380 | 1.65E-41 | 1.49E-40 | -0.08 | 199388 | 91 |
|  |  | Trip Duration | 0.0085 | 199380 | 6.40E-25 | 5.76E-24 | 0.06 | 199388 | 91 |
|  |  | ODI | -0.0068 | 199380 | 0.177 | 1 | -0.07 | 91 | 91 |
|  |  | ESS | 0.0345 | 199380 | 0.016 | 0.140 | 0.12 | 91 | 91 |
|  |  | Age (Yrs) | 0.0196 | 199380 | 0.001 | 0.008 | 0.16 | 91 | 91 |
|  |  | Sex (F) | -0.0292 | 199380 | 0.858 | 1 | -0.01 | 91 | 91 |
|  |  | BMI (kg/m^2^) | 0.0142 | 199380 | 0.213 | 1 | 0.07 | 91 | 91 |
|  | Sig Frequency | Intercept | 0.9133 | 7825 | 8.72E-07 | 7.84E-06 | 2.38 | 7832 | 91 |
|  |  | Trip Duration | -0.0051 | 7825 | 2.58E-25 | 2.32E-24 | -0.25 | 7832 | 91 |
|  |  | ODI | 0.0016 | 7825 | 0.364 | 1 | 0.11 | 91 | 91 |
|  |  | ESS | 0.0002 | 7825 | 0.972 | 1 | 0.00 | 91 | 91 |
|  |  | Age (Yrs) | -0.0040 | 7825 | 0.04997 | 0.450 | -0.25 | 91 | 91 |
|  |  | Sex (F) | -0.0604 | 7825 | 0.292 | 1 | -0.16 | 91 | 91 |
|  |  | BMI (kg/m^2^) | -0.0010 | 7825 | 0.804 | 1 | -0.04 | 91 | 91 |
| Turn | Magnitude | Intercept | 0.3768 | 218934 | 2.13E-20 | 1.91E-19 | 1.91 | 218942 | 91 |
|  |  | Time into Trip | -0.0004 | 218934 | 1.61E-12 | 1.45E-11 | -0.04 | 218942 | 91 |
|  |  | Trip Duration | -0.0002 | 218934 | 0.001 | 0.006 | -0.02 | 218942 | 91 |
|  |  | ODI | -5.41E-05 | 218934 | 0.888 | 1 | -0.01 | 91 | 91 |
|  |  | ESS | 0.0017 | 218934 | 0.116 | 1 | 0.09 | 91 | 91 |
|  |  | Age (Yrs) | -0.0013 | 218934 | 0.003 | 0.028 | -0.16 | 91 | 91 |
|  |  | Sex (F) | 0.0044 | 218934 | 0.719 | 1 | 0.02 | 91 | 91 |
|  |  | BMI (kg/m^2^) | -0.0005 | 218934 | 0.545 | 1 | -0.04 | 91 | 91 |
|  | Duration | Intercept | 7.3294 | 218934 | 3.22E-28 | 2.90E-27 | 1.76 | 218942 | 91 |
|  |  | Time into Trip | -0.0163 | 218934 | 1.52E-36 | 1.37E-35 | -0.07 | 218942 | 91 |
|  |  | Trip Duration | 0.0318 | 218934 | 6.3E-176 | 5.66E-175 | 0.16 | 218942 | 91 |
|  |  | ODI | 0.0041 | 218934 | 0.515 | 1 | 0.03 | 91 | 91 |
|  |  | ESS | 0.0066 | 218934 | 0.708 | 1 | 0.02 | 91 | 91 |
|  |  | Age (Yrs) | -5.64E-05 | 218934 | 0.994 | 1 | -0.0003 | 91 | 91 |
|  |  | Sex (F) | 0.1927 | 218934 | 0.341 | 1 | 0.05 | 91 | 91 |
|  |  | BMI (kg/m^2^) | -0.0266 | 218934 | 0.060 | 0.539 | -0.09 | 91 | 91 |
|  | Sig Frequency | Intercept | 0.8958 | 7825 | 1.61E-06 | 1.45E-05 | 2.18 | 7832 | 91 |
|  |  | Trip Duration | -0.0071 | 7825 | 3.05E-42 | 2.75E-41 | -0.33 | 7832 | 91 |
|  |  | ODI | -0.0003 | 7825 | 0.885 | 1 | -0.02 | 91 | 91 |
|  |  | ESS | 0.0074 | 7825 | 0.137 | 1 | 0.18 | 91 | 91 |
|  |  | Age (Yrs) | -0.0021 | 7825 | 0.304 | 1 | -0.12 | 91 | 91 |
|  |  | Sex (F) | 0.0873 | 7825 | 0.130 | 1 | 0.21 | 91 | 91 |
|  |  | BMI (kg/m^2^) | -0.0042 | 7825 | 0.293 | 1 | -0.14 | 91 | 91 |

#### 3.2 Supplementary Table b: Clinical Classification LME Results

| **Event** | **Feature** | **Predictor** | **Estimate** | **DF** | **P** | **Adjusted**  **P** | **Effect**  **Size (LME)** | **Data Points** | **Pt**  **(n)** |
| --- | --- | --- | --- | --- | --- | --- | --- | --- | --- |
| Brake | Magnitude | Intercept | 0.3860 | 90203 | 1.52E-66 | 1.37E-65 | 3.06 | 11653 | 11 |
|  |  | Time into Trip | -0.0003 | 90203 | 2.57E-08 | 2.32E-07 | -0.05 | 90214 | 91 |
|  |  | Trip Duration | 0.0002 | 90203 | 0.002 | 0.019 | 0.03 | 90214 | 91 |
|  |  | Age (Yrs) | -0.0006 | 90203 | 0.010 | 0.086 | -0.12 | 91 | 91 |
|  |  | Sex (F) | 0.0061 | 90203 | 0.342 | 1 | 0.05 | 91 | 91 |
|  |  | BMI (kg/m^2^) | -0.0007 | 90203 | 0.147 | 1 | -0.07 | 91 | 91 |
|  |  | Non OSA / EDS | 0.0087 | 90203 | 0.420 | 1 | 0.07 | 17548 | 17 |
|  |  | Mild OSA / Non-EDS | 0.0137 | 90203 | 0.226 | 1 | 0.11 | 11868 | 12 |
|  |  | Mild OSA / EDS | 0.0132 | 90203 | 0.237 | 1 | 0.10 | 15773 | 13 |
|  |  | Moderate – Severe / Non-EDS | 0.0204 | 90203 | 0.084 | 0.754 | 0.16 | 8392 | 11 |
|  |  | Moderate – Severe / EDS | 0.0067 | 90203 | 0.527 | 1 | 0.05 | 24980 | 27 |
|  | Duration | Intercept | 5.5694 | 90203 | 7.59E-25 | 6.83E-24 | 1.68 | 11653 | 11 |
|  |  | Time into Trip | -0.0209 | 90203 | 4.84E-38 | 4.36E-37 | -0.11 | 90214 | 91 |
|  |  | Trip Duration | 0.0162 | 90203 | 4.55E-30 | 4.1E-29 | 0.10 | 90214 | 91 |
|  |  | Age (Yrs) | 0.0122 | 90203 | 0.037 | 0.336 | 0.09 | 91 | 91 |
|  |  | Sex (F) | 0.1924 | 90203 | 0.217 | 1 | 0.06 | 91 | 91 |
|  |  | BMI (kg/m^2^) | 0.0052 | 90203 | 0.646 | 1 | 0.02 | 91 | 91 |
|  |  | Non OSA / EDS | -0.1215 | 90203 | 0.643 | 1 | -0.04 | 17548 | 17 |
|  |  | Mild OSA / Non-EDS | 0.0142 | 90203 | 0.959 | 1 | 0.004 | 11868 | 12 |
|  |  | Mild OSA / EDS | 0.5033 | 90203 | 0.063 | 0.568 | 0.15 | 15773 | 13 |
|  |  | Moderate – Severe / Non-EDS | 0.0396 | 90203 | 0.890 | 1 | 0.01 | 8392 | 11 |
|  |  | Moderate – Severe / EDS | 0.2779 | 90203 | 0.281 | 1 | 0.08 | 24980 | 27 |
|  | Sig Frequency | Intercept | 0.7368 | 7822 | 2.01E-06 | 1.81E-05 | 2.53 | 1353 | 11 |
|  |  | Trip Duration | -0.0042 | 7822 | 8.74E-30 | 7.87E-29 | -0.27 | 7832 | 91 |
|  |  | Age (Yrs) | -0.0026 | 7822 | 0.120 | 1 | -0.21 | 91 | 91 |
|  |  | Sex (F) | 0.0544 | 7822 | 0.220 | 1 | 0.19 | 91 | 91 |
|  |  | BMI (kg/m^2^) | -0.0074 | 7822 | 0.020 | 0.184 | -0.36 | 91 | 91 |
|  |  | Non OSA / EDS | 0.1226 | 7822 | 0.095 | 0.854 | 0.42 | 1391 | 17 |
|  |  | Mild OSA / Non-EDS | 0.1616 | 7822 | 0.034 | 0.310 | 0.55 | 880 | 12 |
|  |  | Mild OSA / EDS | 0.0392 | 7822 | 0.605 | 1 | 0.13 | 1391 | 13 |
|  |  | Moderate – Severe / Non-EDS | 0.1671 | 7822 | 0.034 | 0.309 | 0.57 | 692 | 11 |
|  |  | Moderate – Severe / EDS | 0.1519 | 7822 | 0.032 | 0.291 | 0.52 | 2125 | 27 |
| Acceleration | Magnitude | Intercept | 0.2552 | 199377 | 2.77E-25 | 2.5E-24 | 2.18 | 30128 | 11 |
|  |  | Time into Trip | 0.0002 | 199377 | 6.52E-05 | 0.001 | 0.02 | 199388 | 91 |
|  |  | Trip Duration | -0.0006 | 199377 | 4.96E-66 | 4.46E-65 | -0.10 | 199388 | 91 |
|  |  | Age (Yrs) | -0.0008 | 199377 | 0.002 | 0.020 | -0.16 | 91 | 91 |
|  |  | Sex (F) | -0.0042 | 199377 | 0.545 | 1 | -0.04 | 91 | 91 |
|  |  | BMI (kg/m^2^) | -0.0001 | 199377 | 0.768 | 1 | -0.02 | 91 | 91 |
|  |  | Non OSA / EDS | 0.0093 | 199377 | 0.423 | 1 | 0.08 | 34051 | 17 |
|  |  | Mild OSA / Non-EDS | 0.0197 | 199377 | 0.103 | 0.929 | 0.17 | 24676 | 12 |
|  |  | Mild OSA / EDS | 0.0169 | 199377 | 0.159 | 1 | 0.14 | 38608 | 13 |
|  |  | Moderate – Severe / Non-EDS | 0.0217 | 199377 | 0.083 | 0.749 | 0.19 | 17433 | 11 |
|  |  | Moderate – Severe / EDS | 0.0156 | 199377 | 0.166 | 1 | 0.13 | 54492 | 27 |
|  | Duration | Intercept | 3.8645 | 199377 | 4.52E-11 | 4.07E-10 | 1.32 | 30128 | 11 |
|  |  | Time into Trip | -0.0130 | 199377 | 1.76E-41 | 1.58E-40 | -0.08 | 199388 | 91 |
|  |  | Trip Duration | 0.0085 | 199377 | 6.29E-25 | 5.66E-24 | 0.06 | 199388 | 91 |
|  |  | Age (Yrs) | 0.0192 | 199377 | 0.002 | 0.020 | 0.15 | 91 | 91 |
|  |  | Sex (F) | 0.0301 | 199377 | 0.856 | 1 | 0.01 | 91 | 91 |
|  |  | BMI (kg/m^2^) | 0.0071 | 199377 | 0.554 | 1 | 0.03 | 91 | 91 |
|  |  | Non OSA / EDS | 0.4322 | 199377 | 0.120 | 1 | 0.15 | 34051 | 17 |
|  |  | Mild OSA / Non-EDS | 0.3302 | 199377 | 0.252 | 1 | 0.11 | 24676 | 12 |
|  |  | Mild OSA / EDS | 0.6486 | 199377 | 0.024 | 0.215 | 0.22 | 38608 | 13 |
|  |  | Moderate – Severe / Non-EDS | -0.0662 | 199377 | 0.825 | 1 | -0.02 | 17433 | 11 |
|  |  | Moderate – Severe / EDS | 0.5423 | 199377 | 0.044 | 0.397 | 0.18 | 54492 | 27 |
|  | Sig Frequency | Intercept | 0.9705 | 7822 | 1.73E-06 | 1.56E-05 | 2.53 | 1353 | 11 |
|  |  | Trip Duration | -0.0051 | 7822 | 2.67E-25 | 2.4E-24 | -0.25 | 7832 | 91 |
|  |  | Age (Yrs) | -0.0048 | 7822 | 0.027 | 0.240 | -0.30 | 91 | 91 |
|  |  | Sex (F) | -0.0474 | 7822 | 0.414 | 1 | -0.12 | 91 | 91 |
|  |  | BMI (kg/m^2^) | -0.0027 | 7822 | 0.512 | 1 | -0.10 | 91 | 91 |
|  |  | Non OSA / EDS | 0.0047 | 7822 | 0.961 | 1 | 0.01 | 1391 | 17 |
|  |  | Mild OSA / Non-EDS | 0.1465 | 7822 | 0.143 | 1 | 0.38 | 880 | 12 |
|  |  | Mild OSA / EDS | 0.0100 | 7822 | 0.919 | 1 | 0.03 | 1391 | 13 |
|  |  | Moderate – Severe / Non-EDS | 0.0946 | 7822 | 0.360 | 1 | 0.25 | 692 | 11 |
|  |  | Moderate – Severe / EDS | 0.1054 | 7822 | 0.257 | 1 | 0.27 | 2125 | 27 |
| Turn | Magnitude | Intercept | 0.3658 | 218931 | 4.02E-18 | 3.62E-17 | 1.85 | 34901 | 11 |
|  |  | Time into Trip | -0.0004 | 218931 | 1.62E-12 | 1.46E-11 | -0.04 | 218942 | 91 |
|  |  | Trip Duration | -0.0002 | 218931 | 0.001 | 0.006 | -0.02 | 218942 | 91 |
|  |  | Age (Yrs) | -0.0014 | 218931 | 0.002 | 0.022 | -0.16 | 91 | 91 |
|  |  | Sex (F) | 0.0156 | 218931 | 0.189 | 1 | 0.08 | 91 | 91 |
|  |  | BMI (kg/m^2^) | -0.0014 | 218931 | 0.095 | 0.856 | -0.10 | 91 | 91 |
|  |  | Non OSA / EDS | 0.0659 | 218931 | 0.001 | 0.008 | 0.33 | 37887 | 17 |
|  |  | Mild OSA / Non-EDS | 0.0673 | 218931 | 0.001 | 0.010 | 0.34 | 27072 | 12 |
|  |  | Mild OSA / EDS | 0.0625 | 218931 | 0.002 | 0.022 | 0.32 | 43132 | 13 |
|  |  | Moderate – Severe / Non-EDS | 0.0702 | 218931 | 0.001 | 0.010 | 0.35 | 18821 | 11 |
|  |  | Moderate – Severe / EDS | 0.0730 | 218931 | 0.0002 | 0.001 | 0.37 | 57129 | 27 |
|  | Duration | Intercept | 7.5352 | 218931 | 5.00E-25 | 4.5E-24 | 1.81 | 34901 | 11 |
|  |  | Time into Trip | -0.0163 | 218931 | 1.54E-36 | 1.39E-35 | -0.07 | 218942 | 91 |
|  |  | Trip Duration | 0.0318 | 218931 | 6.6E-176 | 5.92E-175 | 0.16 | 218942 | 91 |
|  |  | Age (Yrs) | -0.0021 | 218931 | 0.791 | 1 | -0.01 | 91 | 91 |
|  |  | Sex (F) | 0.2575 | 218931 | 0.210 | 1 | 0.06 | 91 | 91 |
|  |  | BMI (kg/m^2^) | -0.0336 | 218931 | 0.024 | 0.218 | -0.11 | 91 | 91 |
|  |  | Non OSA / EDS | 0.0818 | 218931 | 0.812 | 1 | 0.02 | 37887 | 17 |
|  |  | Mild OSA / Non-EDS | 0.0132 | 218931 | 0.971 | 1 | 0.003 | 27072 | 12 |
|  |  | Mild OSA / EDS | 0.3520 | 218931 | 0.323 | 1 | 0.08 | 43132 | 13 |
|  |  | Moderate – Severe / Non-EDS | 0.3592 | 218931 | 0.333 | 1 | 0.09 | 18821 | 11 |
|  |  | Moderate – Severe / EDS | 0.4599 | 218931 | 0.168 | 1 | 0.11 | 57129 | 27 |
|  | Sig Frequency | Intercept | 0.8916 | 7822 | 4.94E-06 | 4.45E-05 | 2.17 | 1353 | 11 |
|  |  | Trip Duration | -0.0071 | 7822 | 2.81E-42 | 2.53E-41 | -0.33 | 7832 | 91 |
|  |  | Age (Yrs) | -0.0026 | 7822 | 0.220 | 1 | -0.15 | 91 | 91 |
|  |  | Sex (F) | 0.1346 | 7822 | 0.016 | 0.146 | 0.33 | 91 | 91 |
|  |  | BMI (kg/m^2^) | -0.0087 | 7822 | 0.031 | 0.276 | -0.30 | 91 | 91 |
|  |  | Non OSA / EDS | 0.2800 | 7822 | 0.002 | 0.022 | 0.68 | 1391 | 17 |
|  |  | Mild OSA / Non-EDS | 0.2722 | 7822 | 0.005 | 0.042 | 0.66 | 880 | 12 |
|  |  | Mild OSA / EDS | 0.2181 | 7822 | 0.022 | 0.200 | 0.53 | 1391 | 13 |
|  |  | Moderate – Severe / Non-EDS | 0.3053 | 7822 | 0.002 | 0.019 | 0.74 | 692 | 11 |
|  |  | Moderate – Severe / EDS | 0.3158 | 7822 | 0.0004 | 0.004 | 0.77 | 2125 | 27 |

#### 3.3 Supplementary Table c: ODI and ESS Interaction LME Results

| **Event** | **Feature** | **Predictor** | **Estimate** | **DF** | **P** | **Adjusted**  **P** | **Effect**  **Size (LME)** | **Data Points** | **Pt**  **(n)** |
| --- | --- | --- | --- | --- | --- | --- | --- | --- | --- |
| Brake | Magnitude | Intercept | 0.3800 | 90205 | 3.56E-61 | 3.21E-60 | 3.01 | 90214 | 91 |
|  |  | Time into Trip | -0.0003 | 90205 | 2.58E-08 | 2.32E-07 | -0.05 | 90214 | 91 |
|  |  | Trip Duration | 0.0002 | 90205 | 0.002 | 0.019 | 0.03 | 90214 | 91 |
|  |  | Age (Yrs) | -0.0005 | 90205 | 0.027 | 0.243 | -0.10 | 91 | 91 |
|  |  | Sex (F) | 0.0036 | 90205 | 0.562 | 1 | 0.03 | 91 | 91 |
|  |  | BMI (kg/m^2^) | -0.0004 | 90205 | 0.404 | 1 | -0.04 | 91 | 91 |
|  |  | ODI | 0.0004 | 90205 | 0.340 | 1 | 0.10 | 91 | 91 |
|  |  | ESS | 0.0003 | 90205 | 0.693 | 1 | 0.03 | 91 | 91 |
|  |  | ODI*ESS | -0.0001 | 90205 | 0.149 | 1 | -0.06 |  | 91 |
|  | Duration | Intercept | 5.5678 | 90205 | 6.26E-22 | 5.63E-21 | 1.68 | 90214 | 91 |
|  |  | Time into Trip | -0.0209 | 90205 | 4.77E-38 | 4.30E-37 | -0.11 | 90214 | 91 |
|  |  | Trip Duration | 0.0162 | 90205 | 3.95E-30 | 3.55E-29 | 0.10 | 90214 | 91 |
|  |  | Age (Yrs) | 0.0128 | 90205 | 0.028 | 0.248 | 0.09 | 91 | 91 |
|  |  | Sex (F) | 0.1340 | 90205 | 0.392 | 1 | 0.04 | 91 | 91 |
|  |  | BMI (kg/m^2^) | 0.0073 | 90205 | 0.501 | 1 | 0.03 | 91 | 91 |
|  |  | ODI | -0.0094 | 90205 | 0.417 | 1 | -0.08 | 91 | 91 |
|  |  | ESS | 0.0030 | 90205 | 0.880 | 1 | 0.01 | 91 | 91 |
|  |  | ODI*ESS | 0.0008 | 90205 | 0.337 | 1 | 0.04 |  | 91 |
|  | Sig Frequency | Intercept | 0.6815 | 7824 | 4.47E-05 | 0.0004 | 2.34 | 7832 | 91 |
|  |  | Trip Duration | -0.0042 | 7824 | 8.35E-30 | 7.51E-29 | -0.27 | 7832 | 91 |
|  |  | Age (Yrs) | -0.0018 | 7824 | 0.271 | 1 | -0.15 | 91 | 91 |
|  |  | Sex (F) | 0.0361 | 7824 | 0.424 | 1 | 0.12 | 91 | 91 |
|  |  | BMI (kg/m^2^) | -0.0048 | 7824 | 0.126 | 1 | -0.23 | 91 | 91 |
|  |  | ODI | 0.0015 | 7824 | 0.645 | 1 | 0.14 | 91 | 91 |
|  |  | ESS | 0.0038 | 7824 | 0.504 | 1 | 0.13 | 91 | 91 |
|  |  | ODI*ESS | -0.0001 | 7824 | 0.706 | 1 | -0.05 |  | 91 |
| Acceleration | Magnitude | Intercept | 0.2585 | 199379 | 1.14E-23 | 1.02E-22 | 2.20 | 199388 | 91 |
|  |  | Time into Trip | 0.0002 | 199379 | 6.53E-05 | 0.001 | 0.02 | 199388 | 91 |
|  |  | Trip Duration | -0.0006 | 199379 | 5.26E-66 | 4.73E-65 | -0.10 | 199388 | 91 |
|  |  | Age (Yrs) | -0.0008 | 199379 | 0.003 | 0.029 | -0.15 | 91 | 91 |
|  |  | Sex (F) | -0.0073 | 199379 | 0.289 | 1 | -0.06 | 91 | 91 |
|  |  | BMI (kg/m^2^) | 0.0001 | 199379 | 0.867 | 1 | 0.01 | 91 | 91 |
|  |  | ODI | 0.0002 | 199379 | 0.704 | 1 | 0.05 | 91 | 91 |
|  |  | ESS | 0.0001 | 199379 | 0.939 | 1 | 0.01 | 91 | 91 |
|  |  | ODI*ESS | -0.00001 | 199379 | 0.806 | 1 | -0.01 |  | 91 |
|  | Duration | Intercept | 3.6519 | 199379 | 2.75E-09 | 2.48E-08 | 1.24 | 199388 | 91 |
|  |  | Time into Trip | -0.0130 | 199379 | 1.65E-41 | 1.48E-40 | -0.08 | 199388 | 91 |
|  |  | Trip Duration | 0.0085 | 199379 | 6.41E-25 | 5.76E-24 | 0.06 | 199388 | 91 |
|  |  | Age (Yrs) | 0.0197 | 199379 | 0.001 | 0.011 | 0.16 | 91 | 91 |
|  |  | Sex (F) | -0.0279 | 199379 | 0.865 | 1 | -0.01 | 91 | 91 |
|  |  | BMI (kg/m^2^) | 0.0143 | 199379 | 0.212 | 1 | 0.07 | 91 | 91 |
|  |  | ODI | -0.0060 | 199379 | 0.620 | 1 | -0.06 | 91 | 91 |
|  |  | ESS | 0.0356 | 199379 | 0.086 | 0.772 | 0.13 | 91 | 91 |
|  |  | ODI*ESS | -0.0001 | 199379 | 0.941 | 1 | -0.003 |  | 91 |
|  | Sig Frequency | Intercept | 1.0077 | 7824 | 2.16E-06 | 1.95E-05 | 2.63 | 7832 | 91 |
|  |  | Trip Duration | -0.0051 | 7824 | 2.52E-25 | 2.27E-24 | -0.25 | 7832 | 91 |
|  |  | Age (Yrs) | -0.0046 | 7824 | 0.032 | 0.289 | -0.28 | 91 | 91 |
|  |  | Sex (F) | -0.0660 | 7824 | 0.251 | 1 | -0.17 | 91 | 91 |
|  |  | BMI (kg/m^2^) | -0.0013 | 7824 | 0.744 | 1 | -0.05 | 91 | 91 |
|  |  | ODI | -0.0019 | 7824 | 0.660 | 1 | -0.13 | 91 | 91 |
|  |  | ESS | -0.0046 | 7824 | 0.527 | 1 | -0.12 | 91 | 91 |
|  |  | ODI*ESS | 0.0003 | 7824 | 0.367 | 1 | 0.11 |  | 91 |
| Turn | Magnitude | Intercept | 0.3584 | 218933 | 1.09E-14 | 9.79E-14 | 1.81 | 218942 | 91 |
|  |  | Time into Trip | -0.0004 | 218933 | 1.60E-12 | 1.44E-11 | -0.04 | 218942 | 91 |
|  |  | Trip Duration | -0.0002 | 218933 | 0.001 | 0.01 | -0.02 | 218942 | 91 |
|  |  | Age (Yrs) | -0.0012 | 218933 | 0.009 | 0.08 | -0.14 | 91 | 91 |
|  |  | Sex (F) | 0.0056 | 218933 | 0.653 | 1 | 0.03 | 91 | 91 |
|  |  | BMI (kg/m^2^) | -0.0005 | 218933 | 0.595 | 1 | -0.03 | 91 | 91 |
|  |  | ODI | 0.0006 | 218933 | 0.494 | 1 | 0.08 | 91 | 91 |
|  |  | ESS | 0.0026 | 218933 | 0.093 | 0.835 | 0.14 | 91 | 91 |
|  |  | ODI*ESS | -0.0001 | 218933 | 0.413 | 1 | -0.04 |  | 91 |
|  | Duration | Intercept | 7.2832 | 218933 | 1.16E-21 | 1.05E-20 | 1.75 | 218942 | 91 |
|  |  | Time into Trip | -0.0163 | 218933 | 1.52E-36 | 1.37E-35 | -0.07 | 218942 | 91 |
|  |  | Trip Duration | 0.0318 | 218933 | 6.3E-176 | 5.66E-175 | 0.16 | 218942 | 91 |
|  |  | Age (Yrs) | 0.0002 | 218933 | 0.978 | 1 | 0.001 | 91 | 91 |
|  |  | Sex (F) | 0.1955 | 218933 | 0.337 | 1 | 0.05 | 91 | 91 |
|  |  | BMI (kg/m^2^) | -0.0264 | 218933 | 0.062 | 0.562 | -0.09 | 91 | 91 |
|  |  | ODI | 0.0058 | 218933 | 0.700 | 1 | 0.04 | 91 | 91 |
|  |  | ESS | 0.0089 | 218933 | 0.728 | 1 | 0.02 | 91 | 91 |
|  |  | ODI*ESS | -0.0001 | 218933 | 0.901 | 1 | -0.005 |  | 91 |
|  | Sig Frequency | Intercept | 0.7624 | 7824 | 0.0003 | 0.003 | 1.85 | 7832 | 91 |
|  |  | Trip Duration | -0.0071 | 7824 | 3.02E-42 | 2.72E-41 | -0.33 | 7832 | 91 |
|  |  | Age (Yrs) | -0.0013 | 7824 | 0.536 | 1 | -0.08 | 91 | 91 |
|  |  | Sex (F) | 0.0954 | 7824 | 0.097 | 0.877 | 0.23 | 91 | 91 |
|  |  | BMI (kg/m^2^) | -0.0038 | 7824 | 0.342 | 1 | -0.13 | 91 | 91 |
|  |  | ODI | 0.0046 | 7824 | 0.274 | 1 | 0.31 | 91 | 91 |
|  |  | ESS | 0.0141 | 7824 | 0.050 | 0.454 | 0.35 | 91 | 91 |
|  |  | ODI*ESS | -0.0004 | 7824 | 0.204 | 1 | -0.14 |  | 91 |

#### 3.4 Supplementary Table d: Clinical Outcome LME Results

| **Event** | **Feature** | **Predictor** | **Estimate** | **DF** | **P** | **Adjusted**  **P** | **Effect**  **Size (LME)** | **Data Points** | **Pt**  **(n)** |
| --- | --- | --- | --- | --- | --- | --- | --- | --- | --- |
| Brake | Magnitude | Intercept | 0.3837 | 86792 | 2.46E-69 | 2.21E-68 | 3.03 | 40256 | 46 |
|  |  | Time into Trip | -0.0003 | 86792 | 1.24E-06 | 1.11E-05 | -0.04 | 86801 | 88 |
|  |  | Trip Duration | 0.0001 | 86792 | 0.010 | 0.094 | 0.02 | 86801 | 88 |
|  |  | Age (Yrs) | -0.0005 | 86792 | 0.033 | 0.296 | -0.09 | 88 | 88 |
|  |  | Sex (F) | 0.0019 | 86792 | 0.772 | 1 | 0.01 | 88 | 88 |
|  |  | BMI (kg/m^2^) | -0.0005 | 86792 | 0.248 | 1 | -0.06 | 88 | 88 |
|  |  | DC | 0.0025 | 86792 | 0.733 | 1 | 0.02 | 29316 | 23 |
|  |  | FU | -0.0013 | 86792 | 0.878 | 1 | -0.01 | 12877 | 12 |
|  |  | MAD | 0.0122 | 86792 | 0.289 | 1 | 0.10 | 4352 | 7 |
|  | Duration | Intercept | 6.1377 | 86792 | 1.15E-29 | 1.04E-28 | 1.85 | 40256 | 46 |
|  |  | Time into Trip | -0.0199 | 86792 | 1.76E-33 | 1.58E-32 | -0.11 | 86801 | 88 |
|  |  | Trip Duration | 0.0163 | 86792 | 1.89E-29 | 1.70E-28 | 0.10 | 86801 | 88 |
|  |  | Age (Yrs) | 0.0083 | 86792 | 0.146 | 1 | 0.06 | 88 | 88 |
|  |  | Sex (F) | 0.2567 | 86792 | 0.108 | 0.970 | 0.08 | 88 | 88 |
|  |  | BMI (kg/m^2^) | 0.0021 | 86792 | 0.840 | 1 | 0.01 | 88 | 88 |
|  |  | DC | -0.4096 | 86792 | 0.022 | 0.198 | -0.12 | 29316 | 23 |
|  |  | FU | -0.3292 | 86792 | 0.122 | 1 | -0.10 | 12877 | 12 |
|  |  | MAD | -0.2003 | 86792 | 0.484 | 1 | -0.06 | 4352 | 7 |
|  | Sig Frequency | Intercept | 0.6989 | 7524 | 7.14E-06 | 6.43E-05 | 2.39 | 3557 | 46 |
|  |  | Trip Duration | -0.0043 | 7524 | 3.28E-30 | 2.95E-29 | -0.28 | 7532 | 88 |
|  |  | Age (Yrs) | -0.0015 | 7524 | 0.361 | 1 | -0.12 | 88 | 88 |
|  |  | Sex (F) | 0.0186 | 7524 | 0.686 | 1 | 0.06 | 88 | 88 |
|  |  | BMI (kg/m^2^) | -0.0040 | 7524 | 0.196 | 1 | -0.19 | 88 | 88 |
|  |  | DC | 0.0242 | 7524 | 0.634 | 1 | 0.08 | 2428 | 23 |
|  |  | FU | -0.0566 | 7524 | 0.354 | 1 | -0.19 | 1167 | 12 |
|  |  | MAD | 0.0207 | 7524 | 0.801 | 1 | 0.07 | 380 | 7 |
| Acceleration | Magnitude | Intercept | 0.2490 | 192778 | 1.07E-26 | 9.60E-26 | 2.11 | 92686 | 46 |
|  |  | Time into Trip | 0.0002 | 192778 | 2.41E-05 | 0.0002 | 0.02 | 192787 | 88 |
|  |  | Trip Duration | -0.0006 | 192778 | 1.34E-69 | 1.20E-68 | -0.11 | 192787 | 88 |
|  |  | Age (Yrs) | -0.0006 | 192778 | 0.013 | 0.120 | -0.12 | 88 | 88 |
|  |  | Sex (F) | -0.0096 | 192778 | 0.158 | 1 | -0.08 | 88 | 88 |
|  |  | BMI (kg/m^2^) | 0.0002 | 192778 | 0.653 | 1 | 0.02 | 88 | 88 |
|  |  | DC | -0.0006 | 192778 | 0.933 | 1 | -0.01 | 65026 | 23 |
|  |  | FU | 0.0044 | 192778 | 0.629 | 1 | 0.04 | 26131 | 12 |
|  |  | MAD | 0.0172 | 192778 | 0.152 | 1 | 0.15 | 8944 | 7 |
|  | Duration | Intercept | 4.2974 | 192778 | 3.36E-13 | 3.02E-12 | 1.46 | 92686 | 46 |
|  |  | Time into Trip | -0.0127 | 192778 | 3.13E-38 | 2.82E-37 | -0.08 | 192787 | 88 |
|  |  | Trip Duration | 0.0088 | 192778 | 1.69E-25 | 1.52E-24 | 0.06 | 192787 | 88 |
|  |  | Age (Yrs) | 0.0145 | 192778 | 0.018 | 0.158 | 0.12 | 88 | 88 |
|  |  | Sex (F) | -0.0179 | 192778 | 0.918 | 1 | -0.01 | 88 | 88 |
|  |  | BMI (kg/m^2^) | 0.0142 | 192778 | 0.218 | 1 | 0.07 | 88 | 88 |
|  |  | DC | -0.2394 | 192778 | 0.214 | 1 | -0.08 | 65026 | 23 |
|  |  | FU | -0.1787 | 192778 | 0.443 | 1 | -0.06 | 26131 | 12 |
|  |  | MAD | 0.4524 | 192778 | 0.138 | 1 | 0.15 | 8944 | 7 |
|  | Sig Frequency | Intercept | 0.8832 | 7524 | 7.41E-06 | 6.67E-05 | 2.29 | 3557 | 46 |
|  |  | Trip Duration | -0.0053 | 7524 | 2.15E-26 | 1.94E-25 | -0.26 | 7532 | 88 |
|  |  | Age (Yrs) | -0.0029 | 7524 | 0.160 | 1 | -0.18 | 88 | 88 |
|  |  | Sex (F) | -0.0772 | 7524 | 0.185 | 1 | -0.20 | 88 | 88 |
|  |  | BMI (kg/m^2^) | -0.0004 | 7524 | 0.913 | 1 | -0.02 | 88 | 88 |
|  |  | DC | 0.0420 | 7524 | 0.513 | 1 | 0.11 | 2428 | 23 |
|  |  | FU | -0.0633 | 7524 | 0.412 | 1 | -0.16 | 1167 | 12 |
|  |  | MAD | -0.0369 | 7524 | 0.724 | 1 | -0.10 | 380 | 7 |
| Turn | Magnitude | Intercept | 0.4163 | 210142 | 1.18E-21 | 1.06E-20 | 2.10 | 98512 | 46 |
|  |  | Time into Trip | -0.0004 | 210142 | 1.45E-10 | 1.31E-09 | -0.04 | 210151 | 88 |
|  |  | Trip Duration | -0.0002 | 210142 | 0.000 | 0.002 | -0.02 | 210151 | 88 |
|  |  | Age (Yrs) | -0.0015 | 210142 | 0.001 | 0.009 | -0.18 | 88 | 88 |
|  |  | Sex (F) | 0.0054 | 210142 | 0.672 | 1 | 0.03 | 88 | 88 |
|  |  | BMI (kg/m^2^) | -0.0006 | 210142 | 0.468 | 1 | -0.04 | 88 | 88 |
|  |  | DC | -0.0190 | 210142 | 0.182 | 1 | -0.10 | 69077 | 23 |
|  |  | FU | -0.0113 | 210142 | 0.512 | 1 | -0.06 | 33641 | 12 |
|  |  | MAD | 0.0000 | 210142 | 0.999 | 1 | 0.0002 | 8921 | 7 |
|  | Duration | Intercept | 8.0246 | 210142 | 2.87E-29 | 2.58E-28 | 1.93 | 98512 | 46 |
|  |  | Time into Trip | -0.0153 | 210142 | 1.10E-31 | 9.88E-31 | -0.07 | 210151 | 88 |
|  |  | Trip Duration | 0.0319 | 210142 | 3.1E-173 | 2.86E-172 | 0.16 | 210151 | 88 |
|  |  | Age (Yrs) | -0.0055 | 210142 | 0.459 | 1 | -0.03 | 88 | 88 |
|  |  | Sex (F) | 0.2892 | 210142 | 0.167 | 1 | 0.07 | 88 | 88 |
|  |  | BMI (kg/m^2^) | -0.0314 | 210142 | 0.025 | 0.226 | -0.11 | 88 | 88 |
|  |  | DC | -0.4607 | 210142 | 0.048 | 0.432 | -0.11 | 69077 | 23 |
|  |  | FU | -0.1744 | 210142 | 0.536 | 1 | -0.04 | 33641 | 12 |
|  |  | MAD | -0.5229 | 210142 | 0.156 | 1 | -0.13 | 8921 | 7 |
|  | Sig Frequency | Intercept | 1.1047 | 7524 | 4.04E-08 | 3.64E-07 | 2.66 | 3557 | 46 |
|  |  | Trip Duration | -0.0072 | 7524 | 1.16E-40 | 1.04E-39 | -0.33 | 7532 | 88 |
|  |  | Age (Yrs) | -0.0030 | 7524 | 0.146 | 1 | -0.17 | 88 | 88 |
|  |  | Sex (F) | 0.0916 | 7524 | 0.123 | 1 | 0.22 | 88 | 88 |
|  |  | BMI (kg/m^2^) | -0.0053 | 7524 | 0.182 | 1 | -0.18 | 88 | 88 |
|  |  | DC | -0.0719 | 7524 | 0.272 | 1 | -0.17 | 2428 | 23 |
|  |  | FU | -0.0361 | 7524 | 0.647 | 1 | -0.09 | 1167 | 12 |
|  |  | MAD | -0.1262 | 7524 | 0.238 | 1 | -0.30 | 380 | 7 |

#### 3.5 Supplementary Table e: ODI and Time interaction LME Results

| **Event** | **Feature** | **Predictor** | **Estimate** | **DF** | **P** | **Adjusted**  **P** | **Effect**  **Size (LME)** | **Data Points** | **Pt**  **(n)** |
| --- | --- | --- | --- | --- | --- | --- | --- | --- | --- |
| Brake | Magnitude | Intercept | 0.3924 | 90203 | 5.3E-92 | 4.74E-91 | 3.11 | 90214 | 91 |
|  |  | Time into Trip | -0.0013 | 90203 | 2.5E-08 | 2.23E-07 | -0.18 | 90214 | 91 |
|  |  | Trip Duration | 0.0001 | 90203 | 0.119 | 1 | 0.02 | 90214 | 91 |
|  |  | Age (Yrs) | -0.0006 | 90203 | 0.011 | 0.101 | -0.11 | 91 | 91 |
|  |  | Sex (F) | 0.0032 | 90203 | 0.607 | 1 | 0.03 | 91 | 91 |
|  |  | BMI (kg/m^2^) | -0.0005 | 90203 | 0.283 | 1 | -0.05 | 91 | 91 |
|  |  | ODI | -0.00004 | 90203 | 0.865 | 1 | -0.01 | 91 | 91 |
|  |  | Time into Trip*Duration | 0.00003 | 90203 | 0.0002 | 0.002 | 0.04 |  | 91 |
|  |  | Time into Trip*ODI | 0.00001 | 90203 | 0.294 | 1 | 0.02 |  | 91 |
|  |  | Duration*ODI | -0.00001 | 90203 | 0.005 | 0.047 | -0.03 |  | 91 |
|  |  | Time into Trip*Duration*ODI | -0.00000003 | 90203 | 0.932 | 1 | -0.001 |  | 91 |
|  | Duration | Intercept | 5.6777 | 90203 | 1.6E-31 | 1.40E-30 | 1.71 | 90214 | 91 |
|  |  | Time into Trip | -0.0466 | 90203 | 8.3E-15 | 7.43E-14 | -0.25 | 90214 | 91 |
|  |  | Trip Duration | 0.0087 | 90203 | 0.0003 | 0.003 | 0.06 | 90214 | 91 |
|  |  | Age (Yrs) | 0.0134 | 90203 | 0.017 | 0.149 | 0.10 | 91 | 91 |
|  |  | Sex (F) | 0.1261 | 90203 | 0.421 | 1 | 0.04 | 91 | 91 |
|  |  | BMI (kg/m^2^) | 0.0106 | 90203 | 0.327 | 1 | 0.04 | 91 | 91 |
|  |  | ODI | -0.0024 | 90203 | 0.667 | 1 | -0.02 | 91 | 91 |
|  |  | Time into Trip*Duration | 0.0008 | 90203 | 5.2E-06 | 4.70E-05 | 0.05 |  | 91 |
|  |  | Time into Trip*ODI | 0.0003 | 90203 | 0.363 | 1 | 0.02 |  | 91 |
|  |  | Duration*ODI | 0.0001 | 90203 | 0.348 | 1 | 0.01 |  | 91 |
|  |  | Time into Trip*Duration*ODI | -0.00001 | 90203 | 0.433 | 1 | -0.01 |  | 91 |
|  | Sig Frequency | Intercept | 0.7315 | 7825 | 1.1E-07 | 9.99E-07 | 2.51 | 7832 | 91 |
|  |  | Trip Duration | -0.0038 | 7825 | 2.5E-13 | 2.20E-12 | -0.25 | 7832 | 91 |
|  |  | Age (Yrs) | -0.0022 | 7825 | 0.169 | 1 | -0.18 | 91 | 91 |
|  |  | Sex (F) | 0.0315 | 7825 | 0.479 | 1 | 0.11 | 91 | 91 |
|  |  | BMI (kg/m^2^) | -0.0046 | 7825 | 0.136 | 1 | -0.22 | 91 | 91 |
|  |  | ODI | 0.0009 | 7825 | 0.522 | 1 | 0.09 | 91 | 91 |
|  |  | Duration*ODI | -0.00003 | 7825 | 0.280 | 1 | -0.03 |  | 91 |
| Acceleration | Magnitude | Intercept | 0.2604 | 199377 | 2.1E-34 | 1.92E-33 | 2.22 | 199388 | 91 |
|  |  | Time into Trip | -0.0001 | 199377 | 0.441 | 1 | -0.02 | 199388 | 91 |
|  |  | Trip Duration | -0.0005 | 199377 | 8.7E-20 | 7.84E-19 | -0.09 | 199388 | 91 |
|  |  | Age (Yrs) | -0.0008 | 199377 | 0.002 | 0.015 | -0.15 | 91 | 91 |
|  |  | Sex (F) | -0.0074 | 199377 | 0.279 | 1 | -0.06 | 91 | 91 |
|  |  | BMI (kg/m^2^) | 0.0001 | 199377 | 0.893 | 1 | 0.01 | 91 | 91 |
|  |  | ODI | 0.0002 | 199377 | 0.430 | 1 | 0.04 | 91 | 91 |
|  |  | Time into Trip*Duration | 0.00001 | 199377 | 0.246 | 1 | 0.01 |  | 91 |
|  |  | Time into Trip*ODI | 0.000002 | 199377 | 0.839 | 1 | 0.003 |  | 91 |
|  |  | Duration*ODI | -0.00001 | 199377 | 0.004 | 0.033 | -0.02 |  | 91 |
|  |  | Time into Trip*Duration*ODI | 0.0000002 | 199377 | 0.353 | 1 | 0.005 |  | 91 |
|  | Duration | Intercept | 4.1584 | 199377 | 2.1E-15 | 1.88E-14 | 1.42 | 199388 | 91 |
|  |  | Time into Trip | -0.0196 | 199377 | 8.5E-08 | 7.61E-07 | -0.12 | 199388 | 91 |
|  |  | Trip Duration | 0.0060 | 199377 | 2.3E-05 | 0.0002 | 0.04 | 199388 | 91 |
|  |  | Age (Yrs) | 0.0172 | 199377 | 0.004 | 0.036 | 0.14 | 91 | 91 |
|  |  | Sex (F) | -0.0729 | 199377 | 0.664 | 1 | -0.02 | 91 | 91 |
|  |  | BMI (kg/m^2^) | 0.0183 | 199377 | 0.115 | 1 | 0.09 | 91 | 91 |
|  |  | ODI | -0.0062 | 199377 | 0.251 | 1 | -0.06 | 91 | 91 |
|  |  | Time into Trip*Duration | 0.0002 | 199377 | 0.048 | 0.432 | 0.01 |  | 91 |
|  |  | Time into Trip*ODI | -0.0001 | 199377 | 0.454 | 1 | -0.01 |  | 91 |
|  |  | Duration*ODI | 0.00002 | 199377 | 0.752 | 1 | 0.002 |  | 91 |
|  |  | Time into Trip*Duration*ODI | 0.000005 | 199377 | 0.419 | 1 | 0.004 |  | 91 |
|  | Sig Frequency | Intercept | 0.8984 | 7825 | 3.1E-07 | 2.80E-06 | 2.34 | 7832 | 91 |
|  |  | Trip Duration | -0.0042 | 7825 | 6.4E-10 | 5.78E-09 | -0.21 | 7832 | 91 |
|  |  | Age (Yrs) | -0.0040 | 7825 | 0.048 | 0.431 | -0.25 | 91 | 91 |
|  |  | Sex (F) | -0.0600 | 7825 | 0.291 | 1 | -0.16 | 91 | 91 |
|  |  | BMI (kg/m^2^) | -0.0009 | 7825 | 0.810 | 1 | -0.03 | 91 | 91 |
|  |  | ODI | 0.0026 | 7825 | 0.157 | 1 | 0.19 | 91 | 91 |
|  |  | Duration*ODI | -0.0001 | 7825 | 0.080 | 0.724 | -0.04 |  | 91 |
| Turn | Magnitude | Intercept | 0.3935 | 218931 | 5.3E-24 | 4.78E-23 | 1.99 | 218942 | 91 |
|  |  | Time into Trip | -0.0006 | 218931 | 0.013 | 0.121 | -0.05 | 218942 | 91 |
|  |  | Trip Duration | 0.00002 | 218931 | 0.808 | 1 | 0.002 | 218942 | 91 |
|  |  | Age (Yrs) | -0.0014 | 218931 | 0.001 | 0.012 | -0.17 | 91 | 91 |
|  |  | Sex (F) | 0.0024 | 218931 | 0.847 | 1 | 0.01 | 91 | 91 |
|  |  | BMI (kg/m^2^) | -0.0003 | 218931 | 0.723 | 1 | -0.02 | 91 | 91 |
|  |  | ODI | 0.0004 | 218931 | 0.338 | 1 | 0.05 | 91 | 91 |
|  |  | Time into Trip*Duration | 0.000001 | 218931 | 0.870 | 1 | 0.001 |  | 91 |
|  |  | Time into Trip*ODI | -0.00001 | 218931 | 0.302 | 1 | -0.02 |  | 91 |
|  |  | Duration*ODI | -0.00002 | 218931 | 1.5E-05 | 0.0001 | -0.03 |  | 91 |
|  |  | Time into Trip*Duration*ODI | 0.000001 | 218931 | 0.070 | 0.632 | 0.01 |  | 91 |
|  | Duration | Intercept | 7.4563 | 218931 | 2.8E-32 | 2.48E-31 | 1.79 | 218942 | 91 |
|  |  | Time into Trip | -0.0237 | 218931 | 1.3E-06 | 1.18E-05 | -0.10 | 218942 | 91 |
|  |  | Trip Duration | 0.0290 | 218931 | 1.1E-48 | 9.83E-48 | 0.15 | 218942 | 91 |
|  |  | Age (Yrs) | -0.0004 | 218931 | 0.953 | 1 | -0.002 | 91 | 91 |
|  |  | Sex (F) | 0.1845 | 218931 | 0.360 | 1 | 0.04 | 91 | 91 |
|  |  | BMI (kg/m^2^) | -0.0256 | 218931 | 0.067 | 0.604 | -0.09 | 91 | 91 |
|  |  | ODI | 0.0051 | 218931 | 0.442 | 1 | 0.03 | 91 | 91 |
|  |  | Time into Trip*Duration | 0.0003 | 218931 | 0.055 | 0.493 | 0.01 |  | 91 |
|  |  | Time into Trip*ODI | -0.0002 | 218931 | 0.496 | 1 | -0.01 |  | 91 |
|  |  | Duration*ODI | 0.00001 | 218931 | 0.906 | 1 | 0.001 |  | 91 |
|  |  | Time into Trip*Duration*ODI | 0.000003 | 218931 | 0.715 | 1 | 0.002 |  | 91 |
|  | Sig Frequency | Intercept | 0.9770 | 7825 | 5.1E-08 | 4.62E-07 | 2.37 | 7832 | 91 |
|  |  | Trip Duration | -0.0067 | 7825 | 5.2E-20 | 4.71E-19 | -0.31 | 7832 | 91 |
|  |  | Age (Yrs) | -0.0026 | 7825 | 0.199 | 1 | -0.15 | 91 | 91 |
|  |  | Sex (F) | 0.0774 | 7825 | 0.183 | 1 | 0.19 | 91 | 91 |
|  |  | BMI (kg/m^2^) | -0.0032 | 7825 | 0.418 | 1 | -0.11 | 91 | 91 |
|  |  | ODI | 0.0004 | 7825 | 0.826 | 1 | 0.03 | 91 | 91 |
|  |  | Duration*ODI | -0.00003 | 7825 | 0.431 | 1 | -0.02 |  | 91 |

#### 3.6 Supplementary Table f: ESS and Time interaction LME Results

| **Event** | **Feature** | **Predictor** | **Estimate** | **DF** | **P** | **Adjusted**  **P** | **Effect**  **Size (LME)** | **Data Points** | **Pt**  **(n)** |
| --- | --- | --- | --- | --- | --- | --- | --- | --- | --- |
| Brake | Magnitude | Intercept | 0.4136 | 90203 | 4.08E-96 | 3.67E-95 | 3.28 | 90214 | 91 |
|  |  | Time into Trip | -0.0017 | 90203 | 4.55E-05 | 0.0004 | -0.24 | 90214 | 91 |
|  |  | Trip Duration | -0.0003 | 90203 | 0.077 | 0.696 | -0.05 | 90214 | 91 |
|  |  | Age (Yrs) | -0.0006 | 90203 | 0.003 | 0.029 | -0.12 | 91 | 91 |
|  |  | Sex (F) | 0.0031 | 90203 | 0.616 | 1 | 0.02 | 91 | 91 |
|  |  | BMI (kg/m^2^) | -0.0005 | 90203 | 0.183 | 1 | -0.06 | 91 | 91 |
|  |  | ESS | -0.0012 | 90203 | 0.044 | 0.400 | -0.10 | 91 | 91 |
|  |  | Time into Trip*Duration | 3.93E-05 | 90203 | 0.002 | 0.022 | 0.06 |  | 91 |
|  |  | Time into Trip*ESS | 4.87E-05 | 90203 | 0.109 | 0.978 | 0.04 |  | 91 |
|  |  | Duration*ESS | 2.15E-05 | 90203 | 0.093 | 0.840 | 0.02 |  | 91 |
|  |  | Time into Trip*Duration*ESS | -1.11E-06 | 90203 | 0.235 | 1 | -0.01 |  | 91 |
|  | Duration | Intercept | 5.7198 | 90203 | 4.58E-31 | 4.12E-30 | 1.73 | 90214 | 91 |
|  |  | Time into Trip | -0.0442 | 90203 | 7.38E-05 | 0.001 | -0.24 | 90214 | 91 |
|  |  | Trip Duration | -0.0041 | 90203 | 0.375 | 1 | -0.03 | 90214 | 91 |
|  |  | Age (Yrs) | 0.0154 | 90203 | 0.004 | 0.039 | 0.11 | 91 | 91 |
|  |  | Sex (F) | 0.1555 | 90203 | 0.316 | 1 | 0.05 | 91 | 91 |
|  |  | BMI (kg/m^2^) | 0.0089 | 90203 | 0.377 | 1 | 0.04 | 91 | 91 |
|  |  | ESS | -0.0103 | 90203 | 0.494 | 1 | -0.03 | 91 | 91 |
|  |  | Time into Trip*Duration | 0.0006 | 90203 | 0.073 | 0.659 | 0.03 |  | 91 |
|  |  | Time into Trip*ESS | 0.0001 | 90203 | 0.890 | 1 | 0.003 |  | 91 |
|  |  | Duration*ESS | 0.0011 | 90203 | 0.001 | 0.006 | 0.04 |  | 91 |
|  |  | Time into Trip*Duration*ESS | 8.63E-06 | 90203 | 0.724 | 1 | 0.003 |  | 91 |
|  | Sig Frequency | Intercept | 0.7046 | 7825 | 5.72E-07 | 5.15E-06 | 2.42 | 7832 | 91 |
|  |  | Trip Duration | -0.0044 | 7825 | 4.79E-06 | 4.31E-05 | -0.29 | 7832 | 91 |
|  |  | Age (Yrs) | -0.0019 | 7825 | 0.220 | 1 | -0.15 | 91 | 91 |
|  |  | Sex (F) | 0.0332 | 7825 | 0.459 | 1 | 0.11 | 91 | 91 |
|  |  | BMI (kg/m^2^) | -0.0046 | 7825 | 0.117 | 1 | -0.22 | 91 | 91 |
|  |  | ESS | 0.0020 | 7825 | 0.616 | 1 | 0.07 | 91 | 91 |
|  |  | Duration*ESS | 1.81E-05 | 7825 | 0.801 | 1 | 0.01 |  | 91 |
| Acceleration | Magnitude | Intercept | 0.2529 | 199377 | 1.54E-31 | 1.38E-30 | 2.16 | 199388 | 91 |
|  |  | Time into Trip | 0.0002 | 199377 | 0.575 | 1 | 0.02 | 199388 | 91 |
|  |  | Trip Duration | -0.0003 | 199377 | 0.012 | 0.107 | -0.05 | 199388 | 91 |
|  |  | Age (Yrs) | -0.0008 | 199377 | 0.001 | 0.012 | -0.15 | 91 | 91 |
|  |  | Sex (F) | -0.0079 | 199377 | 0.248 | 1 | -0.07 | 91 | 91 |
|  |  | BMI (kg/m^2^) | 0.0001 | 199377 | 0.740 | 1 | 0.02 | 91 | 91 |
|  |  | ESS | 0.0006 | 199377 | 0.358 | 1 | 0.05 | 91 | 91 |
|  |  | Time into Trip*Duration | 1.86E-06 | 199377 | 0.827 | 1 | 0.003 |  | 91 |
|  |  | Time into Trip*ESS | -1.90E-05 | 199377 | 0.343 | 1 | -0.01 |  | 91 |
|  |  | Duration*ESS | -2.77E-05 | 199377 | 0.001 | 0.005 | -0.03 |  | 91 |
|  |  | Time into Trip*Duration*ESS | 4.59E-07 | 199377 | 0.452 | 1 | 0.004 |  | 91 |
|  | Duration | Intercept | 4.1683 | 199377 | 1.48E-15 | 1.33E-14 | 1.42 | 199388 | 91 |
|  |  | Time into Trip | -0.0138 | 199377 | 0.047 | 0.422 | -0.08 | 199388 | 91 |
|  |  | Trip Duration | -0.0095 | 199377 | 0.001 | 0.005 | -0.07 | 199388 | 91 |
|  |  | Age (Yrs) | 0.0180 | 199377 | 0.002 | 0.015 | 0.14 | 91 | 91 |
|  |  | Sex (F) | -0.0070 | 199377 | 0.966 | 1 | -0.002 | 91 | 91 |
|  |  | BMI (kg/m^2^) | 0.0083 | 199377 | 0.435 | 1 | 0.04 | 91 | 91 |
|  |  | ESS | 0.0129 | 199377 | 0.388 | 1 | 0.05 | 91 | 91 |
|  |  | Time into Trip*Duration | 0.0002 | 199377 | 0.246 | 1 | 0.02 |  | 91 |
|  |  | Time into Trip*ESS | -0.0006 | 199377 | 0.228 | 1 | -0.02 |  | 91 |
|  |  | Duration*ESS | 0.0013 | 199377 | 4.98E-10 | 4.48E-09 | 0.05 |  | 91 |
|  |  | Time into Trip*Duration*ESS | 2.38E-06 | 199377 | 0.876 | 1 | 0.001 |  | 91 |
|  | Sig Frequency | Intercept | 0.8267 | 7825 | 4.21E-06 | 3.79E-05 | 2.16 | 7832 | 91 |
|  |  | Trip Duration | -0.0022 | 7825 | 0.078 | 0.699 | -0.11 | 7832 | 91 |
|  |  | Age (Yrs) | -0.0036 | 7825 | 0.067 | 0.607 | -0.22 | 91 | 91 |
|  |  | Sex (F) | -0.0662 | 7825 | 0.246 | 1 | -0.17 | 91 | 91 |
|  |  | BMI (kg/m^2^) | 0.0004 | 7825 | 0.921 | 1 | 0.01 | 91 | 91 |
|  |  | ESS | 0.0041 | 7825 | 0.431 | 1 | 0.11 | 91 | 91 |
|  |  | Duration*ESS | -0.0002 | 7825 | 0.017 | 0.149 | -0.06 |  | 91 |
| Turn | Magnitude | Intercept | 0.3869 | 218931 | 5.53E-23 | 4.98E-22 | 1.96 | 218942 | 91 |
|  |  | Time into Trip | -0.0012 | 218931 | 0.005 | 0.044 | -0.11 | 218942 | 91 |
|  |  | Trip Duration | -0.0008 | 218931 | 1.53E-06 | 1.38E-05 | -0.09 | 218942 | 91 |
|  |  | Age (Yrs) | -0.0013 | 218931 | 0.002 | 0.017 | -0.16 | 91 | 91 |
|  |  | Sex (F) | 0.0046 | 218931 | 0.709 | 1 | 0.02 | 91 | 91 |
|  |  | BMI (kg/m^2^) | -0.0006 | 218931 | 0.476 | 1 | -0.04 | 91 | 91 |
|  |  | ESS | 0.0012 | 218931 | 0.283 | 1 | 0.06 | 91 | 91 |
|  |  | Time into Trip*Duration | 4.74E-05 | 218931 | 0.0003 | 0.002 | 0.04 |  | 91 |
|  |  | Time into Trip*ESS | 3.57E-05 | 218931 | 0.253 | 1 | 0.02 |  | 91 |
|  |  | Duration*ESS | 4.65E-05 | 218931 | 0.0003 | 0.003 | 0.03 |  | 91 |
|  |  | Time into Trip*Duration*ESS | -2.96E-06 | 218931 | 0.002 | 0.017 | -0.01 |  | 91 |
|  | Duration | Intercept | 7.3011 | 218931 | 9.44E-30 | 8.50E-29 | 1.75 | 218942 | 91 |
|  |  | Time into Trip | -0.0327 | 218931 | 0.0003 | 0.002 | -0.14 | 218942 | 91 |
|  |  | Trip Duration | 0.0295 | 218931 | 1.89E-15 | 1.70E-14 | 0.15 | 218942 | 91 |
|  |  | Age (Yrs) | 0.0013 | 218931 | 0.854 | 1 | 0.01 | 91 | 91 |
|  |  | Sex (F) | 0.1827 | 218931 | 0.367 | 1 | 0.04 | 91 | 91 |
|  |  | BMI (kg/m^2^) | -0.0230 | 218931 | 0.081 | 0.726 | -0.08 | 91 | 91 |
|  |  | ESS | 0.0045 | 218931 | 0.806 | 1 | 0.01 | 91 | 91 |
|  |  | Time into Trip*Duration | 0.0004 | 218931 | 0.120 | 1 | 0.02 |  | 91 |
|  |  | Time into Trip*ESS | 0.0005 | 218931 | 0.420 | 1 | 0.01 |  | 91 |
|  |  | Duration*ESS | -2.69E-05 | 218931 | 0.922 | 1 | -0.001 |  | 91 |
|  |  | Time into Trip*Duration*ESS | -8.27E-06 | 218931 | 0.679 | 1 | -0.002 |  | 91 |
|  | Sig Frequency | Intercept | 0.8922 | 7825 | 7.87E-07 | 7.09E-06 | 2.17 | 7832 | 91 |
|  |  | Trip Duration | -0.0064 | 7825 | 2.85E-06 | 2.57E-05 | -0.29 | 7832 | 91 |
|  |  | Age (Yrs) | -0.0022 | 7825 | 0.267 | 1 | -0.13 | 91 | 91 |
|  |  | Sex (F) | 0.0879 | 7825 | 0.126 | 1 | 0.21 | 91 | 91 |
|  |  | BMI (kg/m^2^) | -0.0044 | 7825 | 0.236 | 1 | -0.15 | 91 | 91 |
|  |  | ESS | 0.0083 | 7825 | 0.111 | 1 | 0.21 | 91 | 91 |
|  |  | Duration*ESS | -5.95E-05 | 7825 | 0.557 | 1 | -0.01 |  | 91 |
